# Post-Vaccination Syndrome: A Descriptive Analysis of Reported Symptoms and Patient Experiences After Covid-19 Immunization

**DOI:** 10.1101/2023.11.09.23298266

**Authors:** Harlan M. Krumholz, Yilun Wu, Mitsuaki Sawano, Rishi Shah, Tianna Zhou, Adith S. Arun, Pavan Khosla, Shayaan Kaleem, Anushree Vashist, Bornali Bhattacharjee, Qinglan Ding, Yuan Lu, César Caraballo, Frederick Warner, Chenxi Huang, Jeph Herrin, David Putrino, Danice Hertz, Brianne Dressen, Akiko Iwasaki

**Affiliations:** Center for Outcomes Research and Evaluation, Yale New Haven Hospital, New Haven, Connecticut; Center for Infection and Immunity, Yale School of Medicine, New Haven, Connecticut; Section of Cardiovascular Medicine, Department of Internal Medicine, Yale School of Medicine, New Haven, Connecticut; Department of Health Policy and Management, Yale School of Public Health, New Haven, Connecticut; Department of Biostatistics, Yale School of Public Health, New Haven, Connecticut; Department of Applied Mathematics, Yale College, New Haven, Connecticut; Yale School of Medicine, New Haven, Connecticut; Temerty Faculty of Medicine, University of Toronto, Toronto, Ontario, Canada; The College at the University of Chicago, Chicago, Illinois; Department of Immunobiology, Yale School of Medicine, New Haven, Connecticut; College of Health and Human Sciences, Purdue University, West Lafayette, Indiana; Department of Internal Medicine, Yale School of Medicine, New Haven, Connecticut; Rehabilitation and Human Performance, Icahn School of Medicine at Mount Sinai, New York, New York; Independent Researcher, Los Angeles, California; REACT19; Howard Hughes Medical Institute, Chevy Chase, Maryland

**Author notes:** **Corresponding author:** Dr. Harlan M. Krumholz, 195 Church Street, Fifth Floor, New Haven, CT 06510. Harlan Krumholz and Yilun Wu are co-first authors.

## Abstract

**Introduction:** A chronic post-vaccination syndrome (PVS) after covid-19 vaccination has been reported but has yet to be well characterized.

**Methods:** We included 241 individuals aged 18 and older who self-reported PVS after covid-19 vaccination and who joined the online Yale Listen to Immune, Symptom and Treatment Experiences Now (LISTEN) Study from May 2022 to July 2023. We summarized their demographics, health status, symptoms, treatments tried, and overall experience.

**Results:** The median age of participants was 46 years (interquartile range [IQR]: 38 to 56), with 192 (80%) identifying as female, 209 (87%) as non-Hispanic White, and 211 (88%) from the United States. Among these participants with PVS, 127 (55%) had received the BNT162b2 [Pfizer-BioNTech] vaccine, and 86 (37%) received the mRNA-1273 [Moderna] vaccine. The median time from the day of index vaccination to symptom onset was three days (IQR: 1 day to 8 days). The time from vaccination to symptom survey completion was 595 days (IQR: 417 to 661 days). The median Euro-QoL visual analogue scale score was 50 (IQR: 39 to 70). The five most common symptoms were exercise intolerance (71%), excessive fatigue (69%), numbness (63%), brain fog (63%), and neuropathy (63%). In the week before survey completion, participants reported feeling unease (93%), fearfulness (82%), and overwhelmed by worries (81%), as well as feelings of helplessness (80%), anxiety (76%), depression (76%), hopelessness (72%), and worthlessness (49%) at least once. Participants reported a median of 20 (IQR: 13 to 30) interventions to treat their condition.

**Conclusions:** In this study, individuals who reported PVS after covid-19 vaccination had low health status, high symptom burden, and high psychosocial stress despite trying many treatments. There is a need for continued investigation to understand and treat this condition.

## INTRODUCTION

The vaccines against SARS-CoV-2 have saved many lives, but adverse events have been reported.^1–8^ The Centers for Disease Control and Prevention notes the possibility of rare complications, including anaphylaxis, thrombosis with thrombocytopenia syndrome (specifically after viral vector vaccines), Guillain-Barré Syndrome (specifically after viral vector vaccines), and myocarditis and pericarditis.^9–13^ These complications were not reported in the vaccine clinical trials, emphasizing the limitations of these studies in capturing rare adverse events and highlighting the critical role of post-market surveillance.^14^

A less well-characterized adverse event is a chronic syndrome with symptoms that begin soon after vaccination.^15^ ^16^ A recent preprint from the US National Institutes of Health described 23 people who reported neuropathic symptoms starting within 21 days after vaccination.^17^ The cause of this syndrome is undefined, diagnostic tests and evidence-based interventions are lacking, and its connection with vaccination remains controversial.^18^ A first step in understanding what these patients experience is a description of their symptoms, treatments, and health status. This information can lay the groundwork for enabling prevention, mitigation, and treatment. Accordingly, we sought to describe the characteristics, symptoms, health status, treatment, and experience of individuals who report post-vaccination syndrome (PVS) using data from the Yale Listen to Immune, Symptom and Treatment Experiences Now (LISTEN) study, an online observational study.

## METHODS

### Study Design

LISTEN is a cross-sectional study that collects participant-reported, participant-generated, and clinical data. Deep immune phenotyping is carried out for a subset of individuals, which involves comprehensive profiling of immune cell subtypes, their related products, and their functional states.^19^ LISTEN participants were recruited from Hugo Health Kindred, an online patient community. This platform, for which recruitment occurs mainly through social media and word of mouth, provides opportunities for information exchange, and allows members to share survey responses and health data with research studies.

The LISTEN study began recruiting participants who reported PVS in May 2022. PVS was defined by self-report in response to whether the individual thought the vaccine had injured them. We summarized these individuals’ symptoms, diagnoses, treatments, and experiences.

### Patient Involvement

Patients (DH, BD) were research partners in the study and participated in identifying and prioritizing the research question. Patient partners, including DH, were involved in designing the survey, assessing the burden and time required to participate in the study, recruiting participants, and interpreting results, and will be involved in disseminating the research findings.

### Study Sample

The study sample includes participants aged 18 years and older who reported PVS from May 2022 through July 2023. We did not include people who concurrently reported long covid (**Supplemental Figure 1**).

### Data Collection

The Kindred platform provided a series of surveys (**Supplemental Appendix**) that collected demographic, infection, vaccination, clinical, and social information. The surveys were developed using an iterative process, including feedback from potential participants reporting PVS, to ensure they were relevant and understandable to those participating. The surveys were provided only in English due to funding limitations. Surveys could be completed on computers or mobile devices, and reminders were sent to encourage completion. Surveys were completed between November 2022 and July 2023, with half completed by December 2022 (**Supplemental Figure 2**). Data were extracted on July 7, 2023.

### Variables

The surveys included questions about the participants’ demographic information, health status, and prior SARS-CoV-2 infection and vaccinations. The duration from the day of index vaccination to the day of the survey completion was a median of 595 days (Interquartile Range (IQR): 417 to 661 days; range: 40 to 1058 days).

Prior medical conditions were assessed using the question, “Have you ever been told by a doctor before January 2020 that you have any of the following?” followed by a list of 30 medical diagnostic categories, eight psychiatric diagnostic categories, “other,” and “none of the above.” Current medical conditions were assessed similarly, with 31 medical diagnostic categories, 8 psychiatric diagnostic categories, “other,” and “none of the above.” The question that assessed vaccine-associated symptoms was, “Please select all the following health conditions that you have had as a result of vaccine injury,” followed by a list of 96 specific symptoms, “other,” and “none of the above.” Self-reported health status was assessed on a 5-point scale (excellent, very good, good, fair, or poor) based on individuals’ self-perceived general health. The Euro-QoL visual analogue scale (EQ-VAS) was used to quantify the quality of life, with 100 representing the best.^20^ Symptom severity was collected by asking, “On your worst days, how bad are your symptoms (0 being trivial illness and 100 being unbearable)?”

### Statistical Analysis

We characterized participants by their demographics (age, gender, race, country of residence, marital status, pre-pandemic household income, employment status, and insurance status), vaccine type received, pre-pandemic comorbidities, PVS symptoms, symptom severity, duration of symptoms since vaccination, treatments tried, new-onset medical conditions since vaccination, and psychological and socioeconomic status. These characteristics are described using percentages for categorical variables and median and IQR for continuous variables. We then described the differences in characteristics by age (younger than 60 years vs. 60 years or older), gender, and type of index vaccine. We also summarized the EQ-VAS score on people without prior substantial comorbidities. Such comorbidities did not include common allergies generally perceived as non-life threatening, dyslipidemia, and hypertension. All statistical analyses were performed in R version 4.3.1 (2023-06-16).

The Yale University Institutional Review Board approved the LISTEN study. We followed STROBE reporting guidelines.^21^ Harlan Krumholz, a co-founder of Hugo Health, developed the Hugo Kindred platform, and the Yale Conflict of Interest Committee oversees his involvement.

## RESULTS

### Study Sample

The study population comprised 241 individuals with self-reported PVS (**Supplemental Figure 1**). Among the participants reporting PVS, 127 (55%) were those who had taken the Pfizer-BioNTech vaccine, followed by 86 (37%) who had taken the Moderna vaccine (**Table 1**).

**Table 1.** Vaccinations and Timing of Symptom Onset.

| **Characteristic (N = 241)** | **n** | **%** |
| --- | --- | --- |
| **Number of vaccinations taken** |  |  |
| 1 | 58 | 24 |
| 2 | 87 | 36 |
| 3 | 54 | 22 |
| 4 | 24 | 10 |
| 5 | 13 | 5 |
| Missing | 5 | 2 |
| **Index vaccination type** |  |  |
| AstraZeneca | 4 | 2 |
| Johnson & Johnson | 15 | 6 |
| Moderna | 86 | 37 |
| Novavax | 1 | 0.4 |
| Pfizer | 127 | 55 |
| Missing | 8 | — |
| **Days between index vaccination and onset of any symptoms<sup>1</sup>** | 3 (1-8) | — |
| **Days between index vaccination and onset of severe symptoms<sup>1</sup>** | 6 (2-18) | — |
| **Index vaccination dose** |  |  |
| 1st shot | 106 | 44 |
| 2nd shot | 80 | 33 |
| 3rd shot | 33 | 14 |
| 4th shot | 16 | 7 |
| 5th shot | 6 | 2 |

<sup>1</sup>Median (IQR) for days between index vaccination and onset of any/severe symptoms; n/N, % for all other variables
IQR, Interquartile Range

The participants’ demographic and psychosocial characteristics are shown in **Table 2**. The median age was 46 years (IQR: 38 to 56); 192 (80%) identified as female; 7 (3%) reported that they were non-Hispanic Black and 209 (87%) as non-Hispanic White. There were 127 (55%) participants who received the BNT162b2 [Pfizer-BioNTech] vaccine, 86 (37%) the mRNA-1273 [Moderna] vaccine, 15 (6%) the Janssen Ad26.COV2.S [Johnson & Johnson] vaccine, 1 (0.4%) the NVX-CoV2373 [Novavax] vaccine, and 4 (2%) the ChAdOx1 nCoV-19 [AstraZeneca] vaccine. Additionally, 82 (34%) participants reported being infected by the SARS-CoV-2 virus at least once.

**Table 2.**
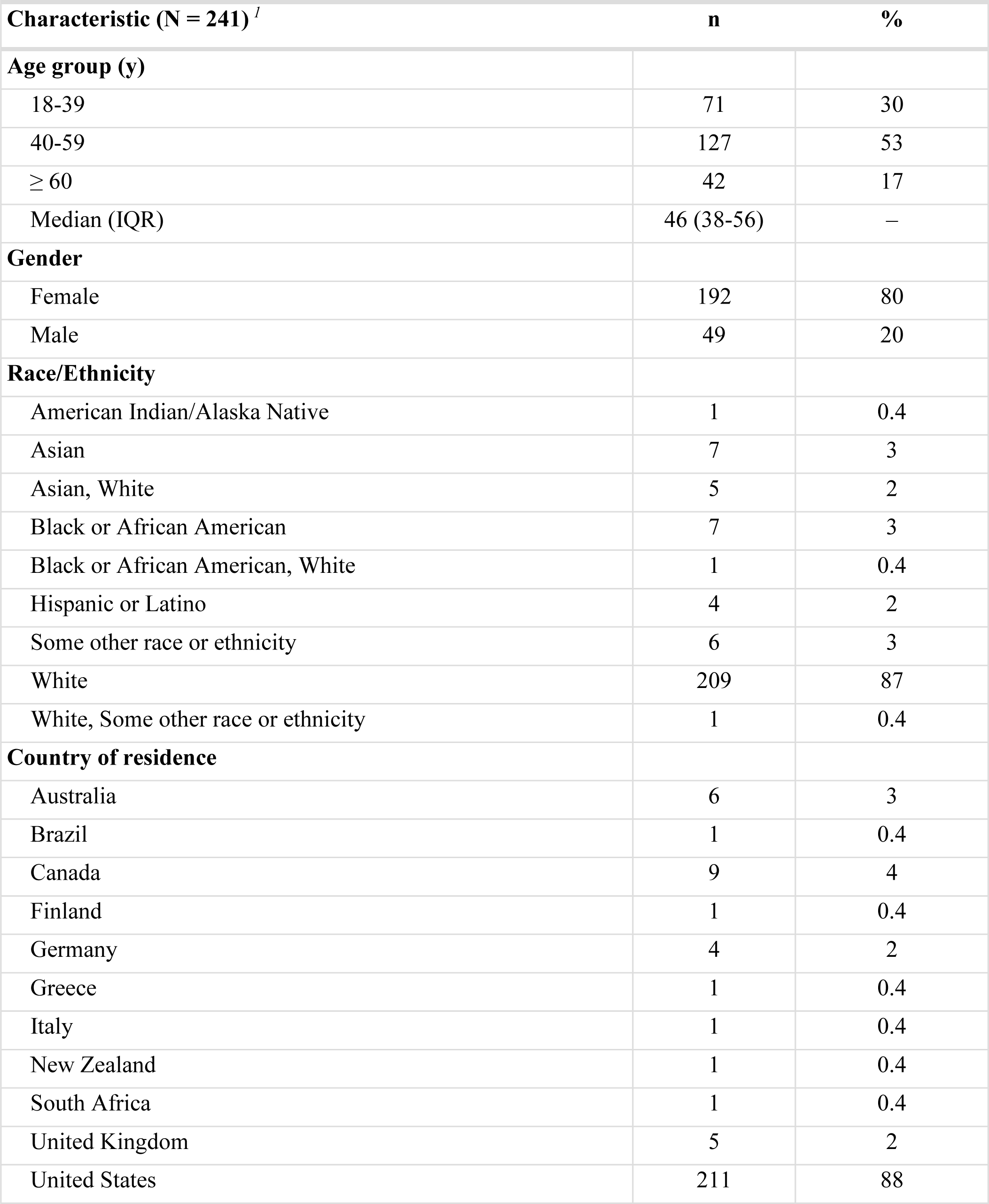
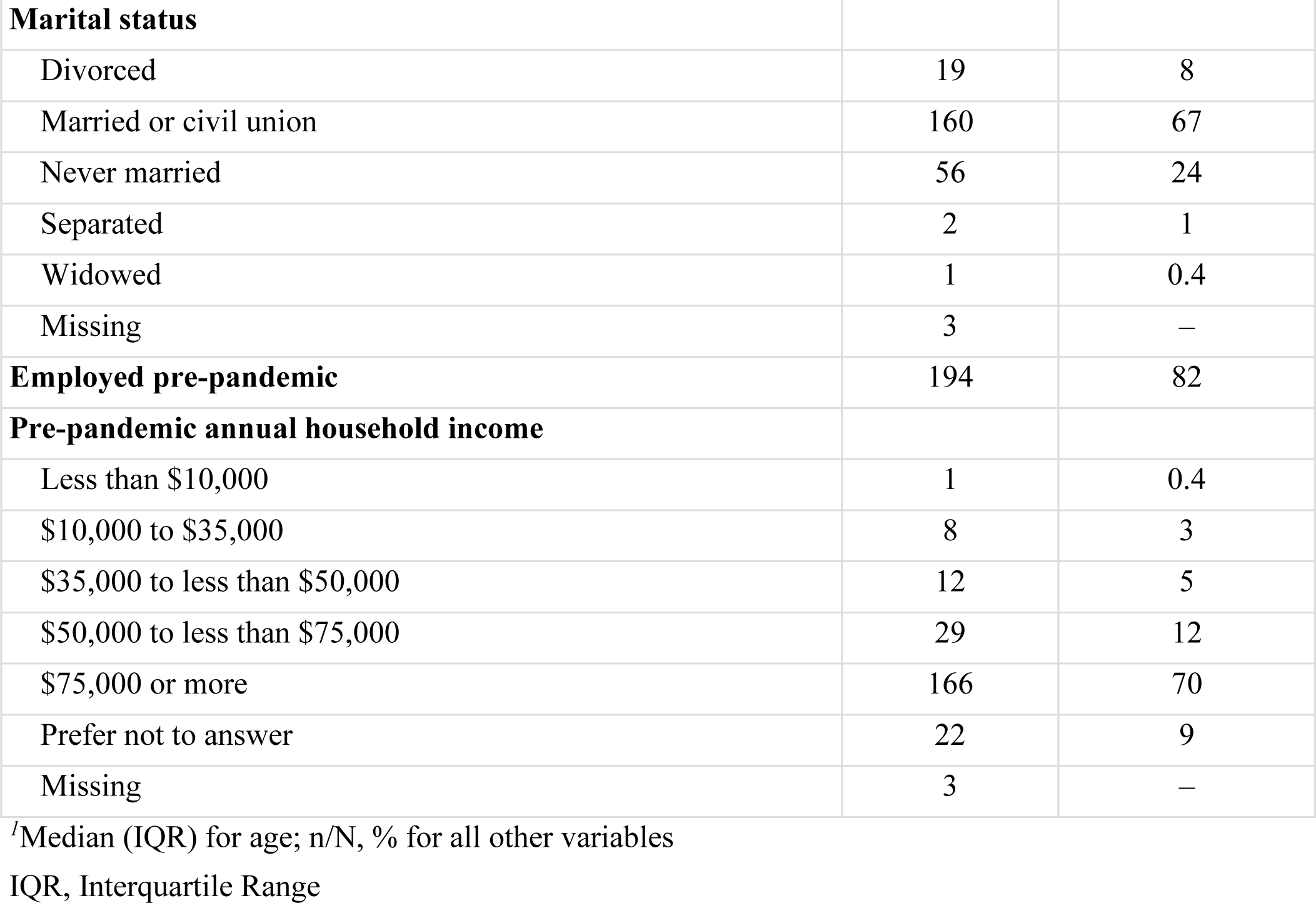
Participant Demographic and Psychosocial Characteristics for Those Reporting PVS.

| **Characteristic (N = 241) <sup>1</sup>** | **n** | **%** |
| --- | --- | --- |
| **Age group (y)** |  |  |
| 18-39 | 71 | 30 |
| 40-59 | 127 | 53 |
| ≥ 60 | 42 | 17 |
| Median (IQR) | 46 (38-56) | – |
| **Gender** |  |  |
| Female | 192 | 80 |
| Male | 49 | 20 |
| **Race/Ethnicity** |  |  |
| American Indian/Alaska Native | 1 | 0.4 |
| Asian | 7 | 3 |
| Asian, White | 5 | 2 |
| Black or African American | 7 | 3 |
| Black or African American, White | 1 | 0.4 |
| Hispanic or Latino | 4 | 2 |
| Some other race or ethnicity | 6 | 3 |
| White | 209 | 87 |
| White, Some other race or ethnicity | 1 | 0.4 |
| **Country of residence** |  |  |
| Australia | 6 | 3 |
| Brazil | 1 | 0.4 |
| Canada | 9 | 4 |
| Finland | 1 | 0.4 |
| Germany | 4 | 2 |
| Greece | 1 | 0.4 |
| Italy | 1 | 0.4 |
| New Zealand | 1 | 0.4 |
| South Africa | 1 | 0.4 |
| United Kingdom | 5 | 2 |
| United States | 211 | 88 |
| **Marital status** |  |  |
| Divorced | 19 | 8 |
| Married or civil union | 160 | 67 |
| Never married | 56 | 24 |
| Separated | 2 | 1 |
| Widowed | 1 | 0.4 |
| Missing | 3 | – |
| **Employed pre-pandemic** | 194 | 82 |
| **Pre-pandemic annual household income** |  |  |
| Less than \$10,000 | 1 | 0.4 |
| \$10,000 to \$35,000 | 8 | 3 |
| \$35,000 to less than \$50,000 | 12 | 5 |
| \$50,000 to less than \$75,000 | 29 | 12 |
| \$75,000 or more | 166 | 70 |
| Prefer not to answer | 22 | 9 |
| Missing | 3 | – |

<sup>1</sup>Median (IQR) for age; n/N, % for all other variables
IQR, Interquartile Range

The participants were mainly from the United States (88%), and in this group, there were 5 uninsured individuals, 176 with private commercial insurance, and 8 Medicaid beneficiaries. Before the pandemic, 194 (82%) were employed. Regarding income, 166 (69%) reported an annual income greater than or equal to $75,000. Approximately three quarters had one or more pre-pandemic comorbidities including gastrointestinal issues reported by 67 (28%), anxiety by 61 (25%), asthma by 49 (20%), depression by 49 (20%), and migraines by 46 (19%). There were 59 (25%) participants who did not have any prior substantial comorbidities. The frequencies of pre-pandemic comorbidities are shown in **Supplemental Table 1**.

The duration from the day of index vaccination to the day of the survey completion was a median of 595 days (IQR: 417 to 661 days; range: 40 to 1058 days).

Participants reported numerous challenges in their daily lives (**Supplemental Table 2**). In the week before survey completion, 221 (93%) reported feeling unease at least once, 194 (82%) felt fearful, 192 (81%) felt overwhelmed by worries, and 180 (76%) struggled with anxiety. Furthermore, 190 (80%) felt helpless, 182 (76%) depressed, 171 (72%) hopeless, and 116 (49%) worthless at least once in the week before survey completion. Additionally, 233 (98%) felt rundown and 216 (91%) reported sleep problems. Pain interfered with the daily activities of 204 (86%) participants.

Regarding social support, 98 (41%) had two or fewer supportive individuals to rely on for help. Getting help from neighbors was described as challenging or very challenging by 86 (36%), while 24 (10%) rarely or never had assistance for tasks like shopping or visiting the doctor. Loneliness was prevalent, with 47 (20%) often feeling a lack of companionship, 55 (23%) feeling left out, and 77 (32%) feeling isolated. Furthermore, 28 (12%) often or always felt lonely.

Concerns related to living situations and food security were also prominent. Among all participants, 21 (9%) feared running out of food before they could afford to buy more, and 16 (7%) had stable housing but expressed worry about future loss. Transportation issues impeded 13 (5%) participants from carrying out essential non-medical tasks and 10 (4%) from attending medical appointments.

### Timing of Symptom Onset Following Vaccination

The median time from vaccination to the onset of any symptoms was 3 days (IQR: 1 to 8 days) (**Table 2**; **Supplemental Figure 3**). Symptoms began after the first, second, third, and fourth (or more) vaccinations for 106 (44%), 80 (33%), 33 (14%), and 22 (9%) participants, respectively.

### Health Status

The median EQ-VAS score among participants was 50 (IQR: 39 to 70) (**Figure 1**). We evaluated the distributions of EQ-VAS scores in men (median: 50; IQR: 37 to 70) and women (median: 51; IQR: 39 to 69), age groups (<60 years old median: 50; IQR: 39 to 65; ≥60 years old median: 61; IQR: 39 to 76)), and Pfizer-BioNTech (median: 51; IQR: 35 to 70) and Moderna (median: 50; IQR: 40 to 62) index vaccination types (**Figure 1**). The median EQ-VAS score in the participants without previous comorbidities was 52 (IQR: 36 to 70). The median EQ-VAS score in the participants with any prior comorbidities was 50 (IQR: 39 to 66). There were 106 (44%) participants who rated their current health as fair or poor (**Supplemental Table 3**; **Supplemental Figure 4**), and their median EQ-VAS score was 40 (IQR: 30 to 52).

**Figure 1.**
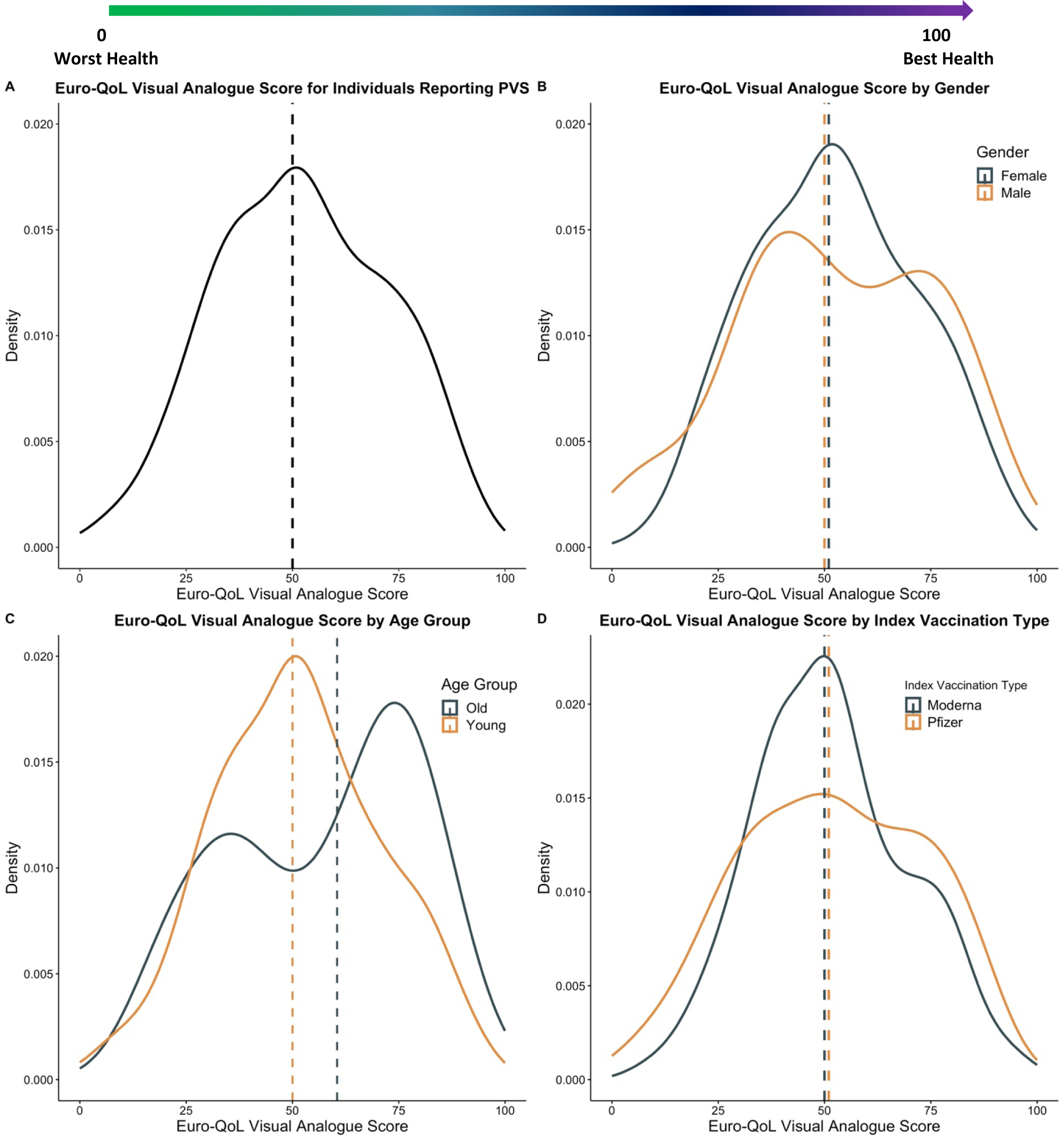
Distribution of health status, measured by the Euro-QoL visual analogue scale. Presented for (A) all participants who reported PVS, (B) all participants who reported PVS stratified by gender, (C) all participants who reported PVS stratified by age group (old if aged 60 years or above), and (D) all participants who reported PVS stratified by index vaccination type. Median values are indicated with dashed lines. Assessed by the question, “Please choose one point in this 0-100 scale, which can best represent your health today (0 means the worst and 100 means the best).” (**Supplemental Appendix** [Health status questions]). IQR, Interquartile Range; PVS, Post-vaccination Syndrome

### Symptom Severity

When asked to quantify symptom severity on their worst days (0 representing a trivial illness and 100 for an unbearable condition), participants reported a median severity of 80 (IQR: 69 to 89, **Supplemental Figure 5**; **Supplemental Table 3**). On their worst days, participants who rated their current health as fair or poor reported a median symptom severity of 80 points (IQR: 70 to 90).

### Symptom Characteristics

The symptoms reported by the participants are shown in **Figure 2**, **Supplemental Figure 6**, and **Supplemental Table 4**. The median number of symptoms attributed to PVS was 22 (IQR: 13 to 35). The most common symptoms were exercise intolerance reported by 170 (71%) participants, excessive fatigue by 167 (69%), numbness by 153 (63%), brain fog by 151 (63%), neuropathy by 151 (63%), insomnia by 148 (61%), palpitations by 145 (60%), myalgia by 132 (55%), tinnitus or humming in ears by 131 (54%), headache by 128 (53%), burning sensations by 121 (50%), and dizziness by 121 (50%).

**Figure 2.**
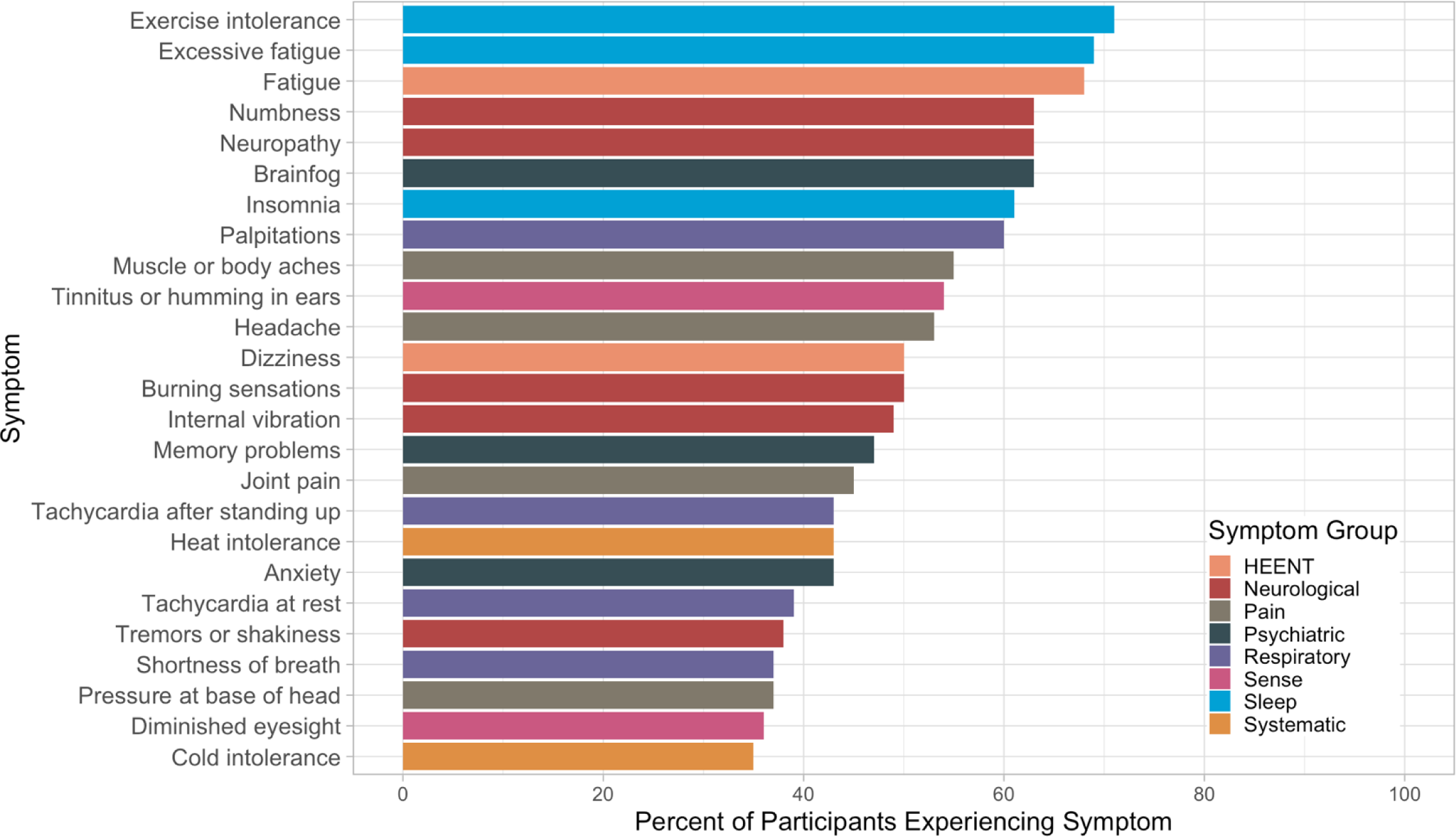
Top 25 most common symptoms with their corresponding symptom groups within 241 participants who reported PVS. HEENT, Head, Ears, Eyes, Nose, Throat; PVS, Post-vaccination Syndrome

### New Diagnoses Since the Pandemic

The most common new diagnoses in the study sample since the beginning of the pandemic were anxiety (49 (36%) participants), neurological conditions (79 [33%]), gastrointestinal issues (73 [30%]), and postural orthostatic tachycardia syndrome (POTS) (70 [29%]) (**Supplemental Table 5**). There were 53 (22%) participants who reported migraine and 49 (20%) who reported depression.

### Treatments

Participants reported having taken many treatments (**Figure 3**, **Figure 4**, **Supplemental Table 6**). The total number of unique treatment types was 209, which we grouped into 40 categories. The median number of individual treatments tried was 20 (IQR: 13 to 30; range: 0 to 65). The most common prescription therapies reported were oral steroids (116 (48%) participants), gabapentin (61 [25%]), low-dose naltrexone (48 [20%]), ivermectin (44 [18%]), propranolol (27 [11%]), and bronchodilators (26 [11%]). More than 500 additional treatments were reported by participants (**Supplemental Table 6**). The most common non-pharmacological treatment included limiting exercise or exertion by 124 (51%) participants, quitting alcohol or caffeine (105 [44%]), hydration and increasing salt intake (105 [44%]), and intermittent fasting (95 [39%]).

**Figure 3.**
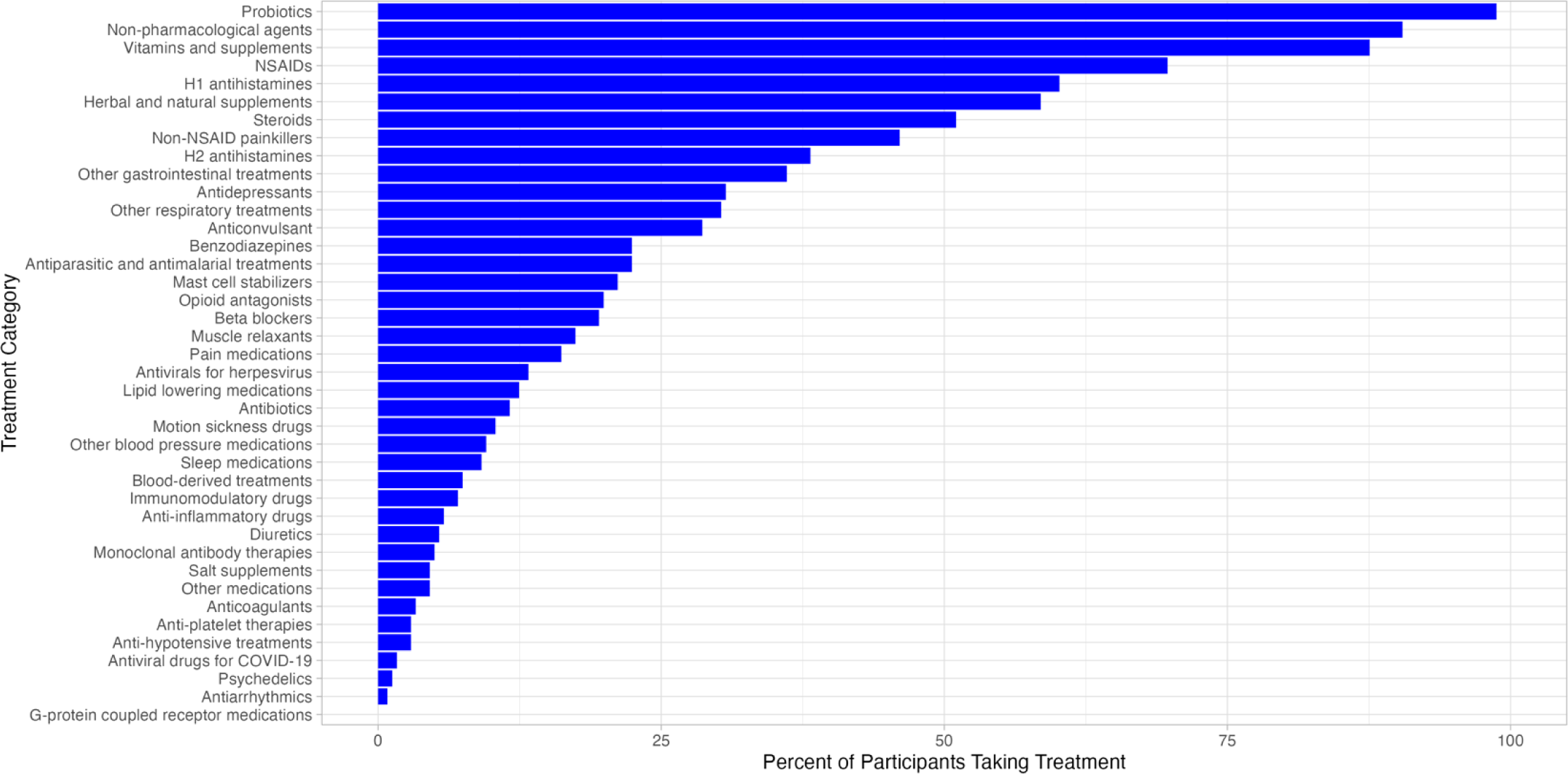
The frequency of treatments tried among PVS participants, grouped by categories. NSAID, Non-steroidal Anti-inflammatory Drug; PVS, Post-vaccination Syndrome

**Figure 4.**
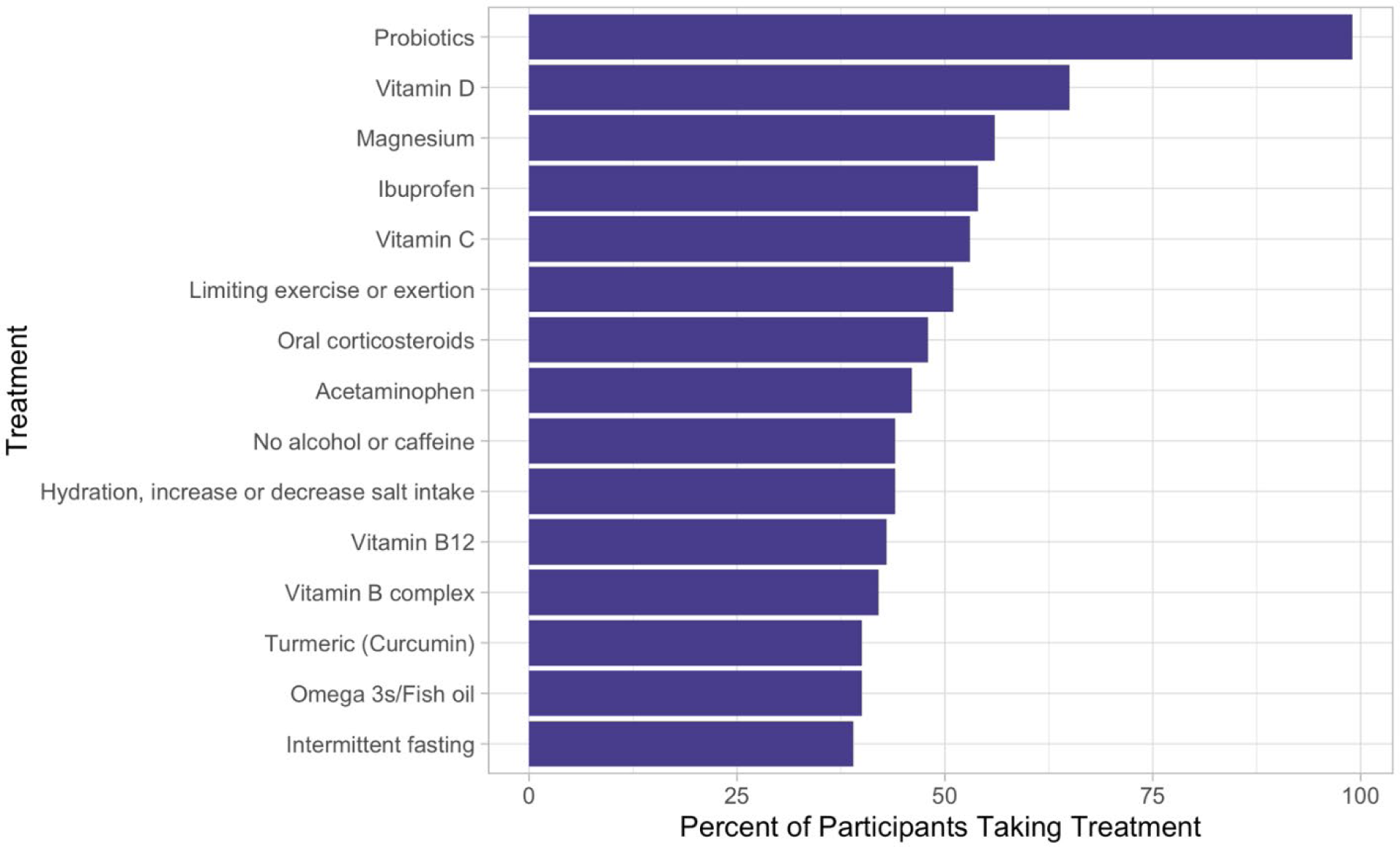
Top 15 most-tried individual treatments among PVS participants. The “probiotics” category contains probiotics as the only component. PVS, Post-vaccination Syndrome

## DISCUSSION

This study is the largest ^5^ ^15^ ^22–27^ to describe people who report a severe, debilitating chronic condition following covid-19 vaccination. This chronic condition began soon after covid-19 vaccination and persisted in many people for a year or more. The symptoms reported are diverse and severe. Despite having tried many treatments, the median EQ-VAS score was low.^20^ Although the cause of PVS has yet to be established, this report is a step toward acknowledging the suffering it creates. It signals the need for scientific investigation to help us understand its mechanism and develop prevention, mitigation, and cure strategies.

This observational study of self-referred individuals cannot determine causality or provide estimates of the incidence and prevalence of PVS. Although there is a background rate for conditions unrelated to vaccination that can produce many symptoms reported by participants, these individuals do not have other diagnoses to explain their symptoms.^28^ Many participants did not have chronic conditions before the pandemic. Also, we excluded LISTEN participants who reported long covid.

PVS could be caused by several potential mechanisms, including a mechanism related to the vaccination or manufacturing process.^1^ It may represent a rare response to vaccines in susceptible individuals.^29^ ^30^ Some investigators have concluded, based on their self-referred case series, that vaccines may cause immune system dysfunction.^17^ They focused on people with neuropathy after vaccination and could not identify other causes. In 2 of 5 participants, the cerebrospinal fluid had oligoclonal bands, and they found immune complexes in some skin biopsies. Nevertheless, the syndrome could be unrelated to the vaccination, occurring by chance during the vaccination period.^2^ ^31^ ^32^ However, the temporal relationship with clustering of symptom onset within the first 1–18 days from the index vaccine suggests a potential relationship. The possibility that the syndrome may be related to the vaccination has implications for future vaccine development and safety surveillance.^33–35^

Research in this area has the risk of being embroiled in debates about vaccinations. The net benefit of the covid-19 vaccination program is clear,^1^ ^13^ ^36–38^ and there are concerns about vaccine hesitancy. But fears of inciting vaccine hesitancy^39^ ^40^ should not impede efforts to research this condition—and make progress for people who are suffering.

This study has several limitations. It is an uncontrolled observational cross-sectional study of self-referred participants who reported that they had the onset of symptoms soon after vaccination, which, in many cases, persisted for more than a year. The participants are not representative, so it is not possible to estimate the incidence or who might be most susceptible to this condition. Participating in LISTEN required people to join the community, consent to the study, and complete the surveys. This approach may have skewed our sample away from people who were too ill to participate or had substantial cognitive dysfunction. Also, the study required online access, some digital literacy, and English fluency, further limiting participation and representativeness. Since participants were strongly skewed toward those who reside in the United States and self-identify as White, efforts are needed to study a more inclusive group. Future investigations need more outreach and partnerships with diverse groups, low-income communities, communities of color, and other countries to assemble a more representative participant group.

## CONCLUSION

In conclusion, people reporting PVS after covid-19 vaccination in this study are highly symptomatic, have poor health status, and have tried many treatment strategies without success. As PVS is associated with considerable suffering, there is an urgent need to understand its mechanism to provide prevention, diagnosis, and treatment strategies.

## Supporting information

Supplemental Protocol

STROBE Checklist

## Data Availability

Deidentified data in the present study are available upon reasonable request to the authors.

## Conflict of Interest Disclosures

Harlan Krumholz received expenses and/or personal fees from Element Science, Eyedentify, and F-Prime in the past three years. He is a co-founder of Refactor Health and Ensight-AI. He and his spouse are co-founders of, and have equity in, Hugo Health, the personalized health data platform company that developed the Hugo Kindred platform. His spouse is an officer with Hugo Health. The Yale Conflict of Interest Committee oversees his involvement in this study. He is the editor of Journal Watch: Cardiology of the Massachusetts Medical Society and a section editor for UpToDate. He is associated with contracts, through Yale New Haven Hospital, from the Centers for Medicare & Medicaid Services, and through Yale University from Janssen, Johnson & Johnson Consumer, and Pfizer. Akiko Iwasaki co-founded RIGImmune, Xanadu Bio, and PanV, consults for Paratus Sciences and InvisiShield Technologies, and is a member of the Board of Directors of Roche Holding Ltd. Yuan Lu received research grants from the United States National Institutes of Health, the Patient-Centered Outcomes Research Institute, and the Sentara Research Foundation outside of the submitted work. Jeph Herrin receives funding from multiple institutes of the National Institutes of Health, from the Patient-Centered Outcomes Research Institute, the American Heart Association, and the Agency for Healthcare Research and Quality for research projects; from the Centers for Medicare & Medicaid Services for development of quality measures; and from Pfizer. Chenxi Huang receives K12 funding from the National Center for Advancing Translational Science of the National Institutes of Health (UL1TR001863). Bornali Bhattacharjee is supported by, and César Caraballo was supported by, a grant from the Yale-Mayo Clinic Center of Excellence in Regulatory Science and Innovation (CERSI) (U01FD005938). The other authors have no financial relationships to disclose.

## Funding

This project was in part supported by the Howard Hughes Medical Institute Collaborative COVID-19 Initiative and in part supported by CTSA Grant Number UL1 TR001863 from the National Center for Advancing Translational Science, a component of the National Institutes of Health. Its contents are solely the responsibility of the authors and do not necessarily represent the official views of the National Institutes of Health.

## Notes

### Author Declarations

The Yale University Institutional Review Board gave ethical approval to the LISTEN (Listen to Immune, Symptom and Treatment Experiences Now) study. STROBE reporting guidelines were followed. Harlan Krumholz, a co-founder of Hugo Health, developed the Hugo Kindred platform, and the Yale Conflict of Interest Committee oversees his involvement.

