## Supplemental Figures and Tables for "Post-Vaccination Syndrome: A Descriptive Analysis of Reported Symptoms and Patient Experiences After Covid-19 Immunization"

**TABLE OF CONTENTS**
**Supplemental Figures and Tables**

| Supplemental Document | Page |
|---|---|
| Supplemental Figure 1. Sample selection………………………………………………………… | 2 |
| Supplemental Figure 2. Survey completion time………………………………………………… | 3 |
| Supplemental Figure 3. Cumulative distribution of days between index vaccination and onset of (A) any symptoms and (B) severe symptoms ………………………………………………. | 4 |
| Supplemental Figure 4. Self-reported health status ……………………………………….…… | 5 |

**Supplemental Figure 1. Sample selection.**

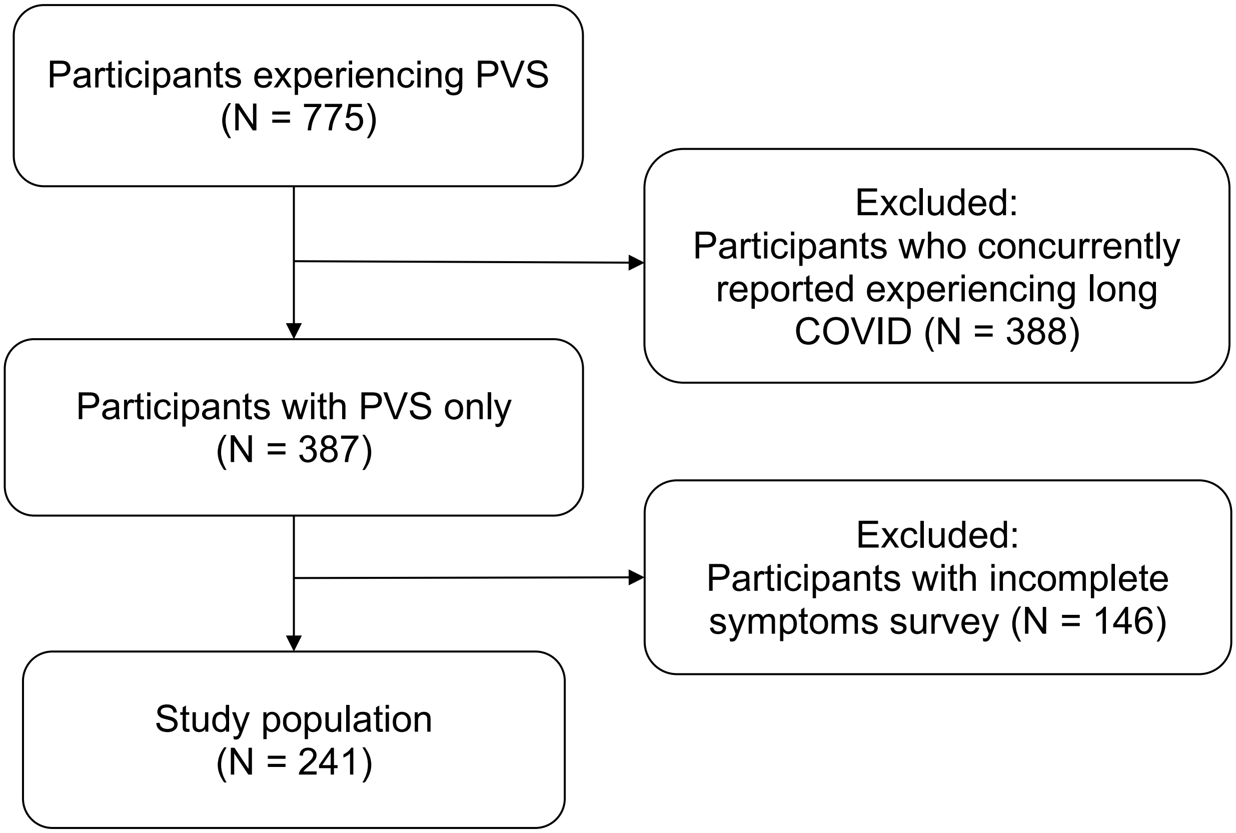

LISTEN, Listen to Immune, Symptom and Treatment Experiences Now; PVS, Post-vaccination Syndrome

**Supplemental Figure 2. Survey completion time.**

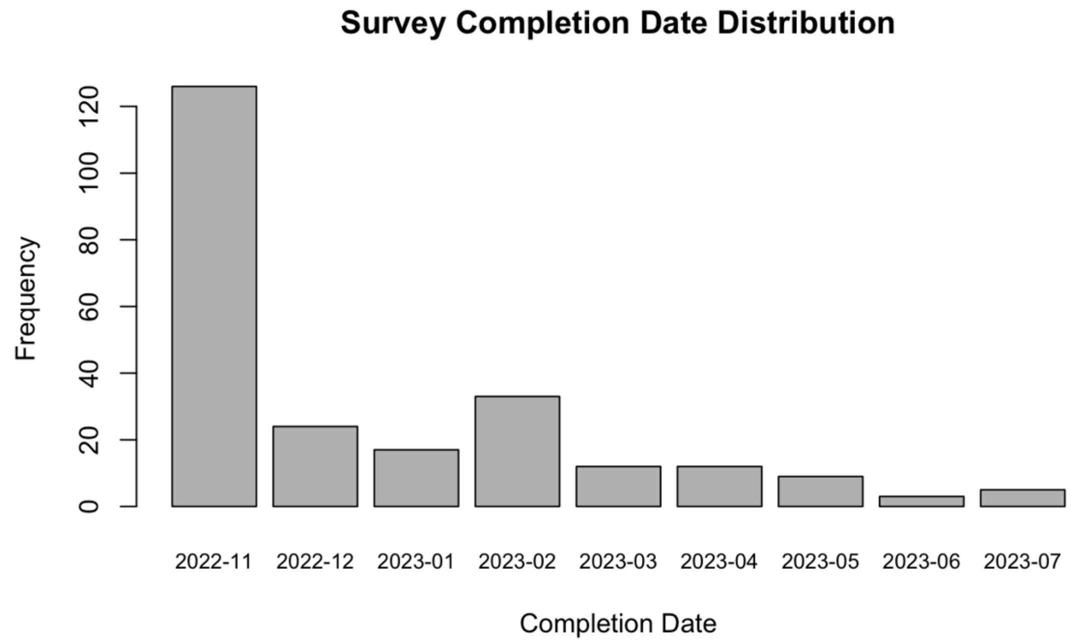

**Supplemental Figure 3. Cumulative distribution of days between index vaccination and onset of (A) any symptoms and (B) severe symptoms.**

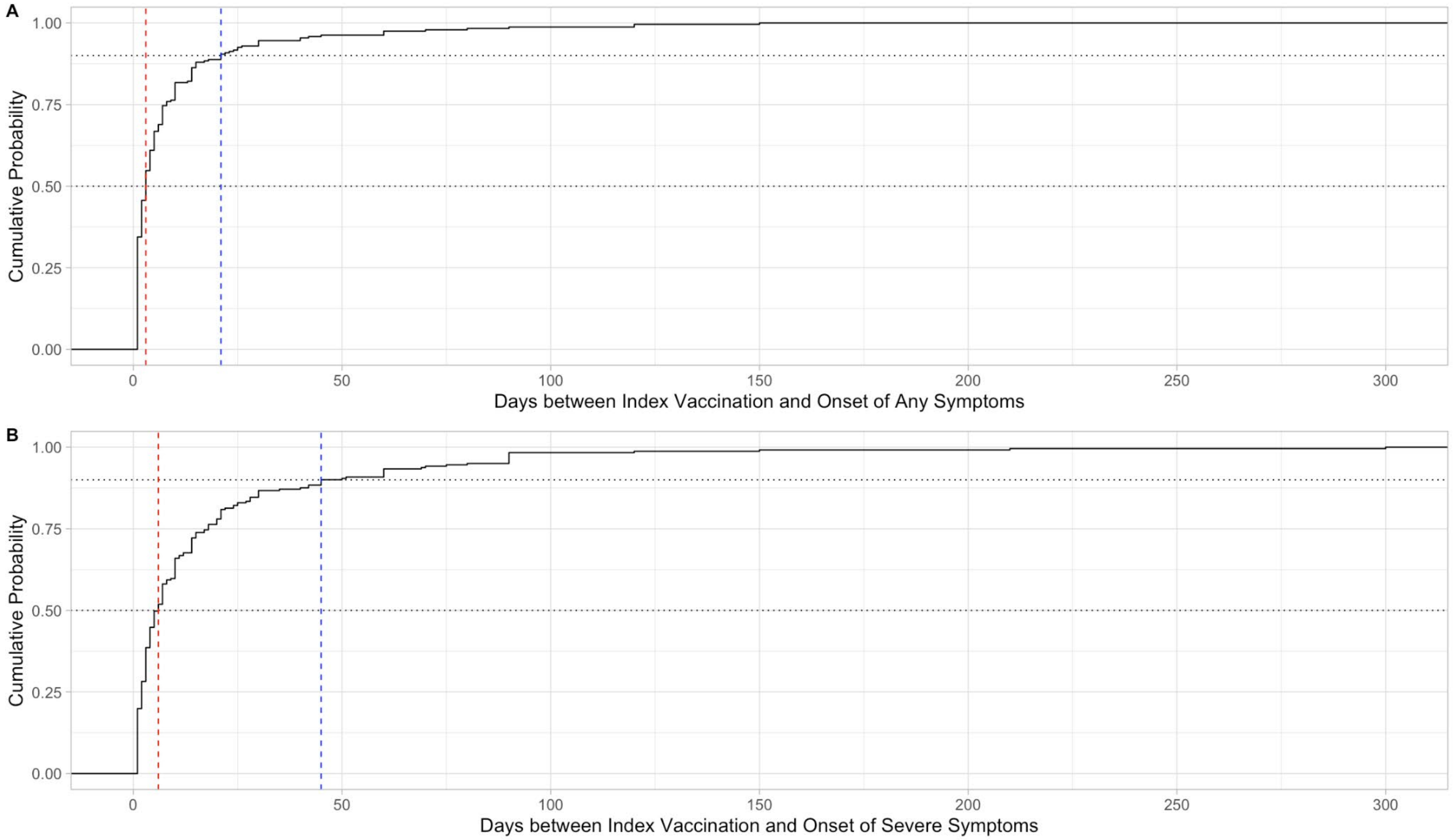

The red line indicates the median and the blue line indicates the 90% percentile.
Half of PVS participants developed any symptoms 3 days after their index vaccination and 90% developed any symptoms after 21 days of their index vaccination.
Half of PVS participants developed severe symptoms 6 days after their index vaccination and 90% developed severe symptoms 45 days after their index vaccination.
The black dotted line represents a cumulative probability of 0.5 and 0.9.
PVS, Post-vaccination Syndrome

**Supplemental Figure 4. Self-reported health status**.

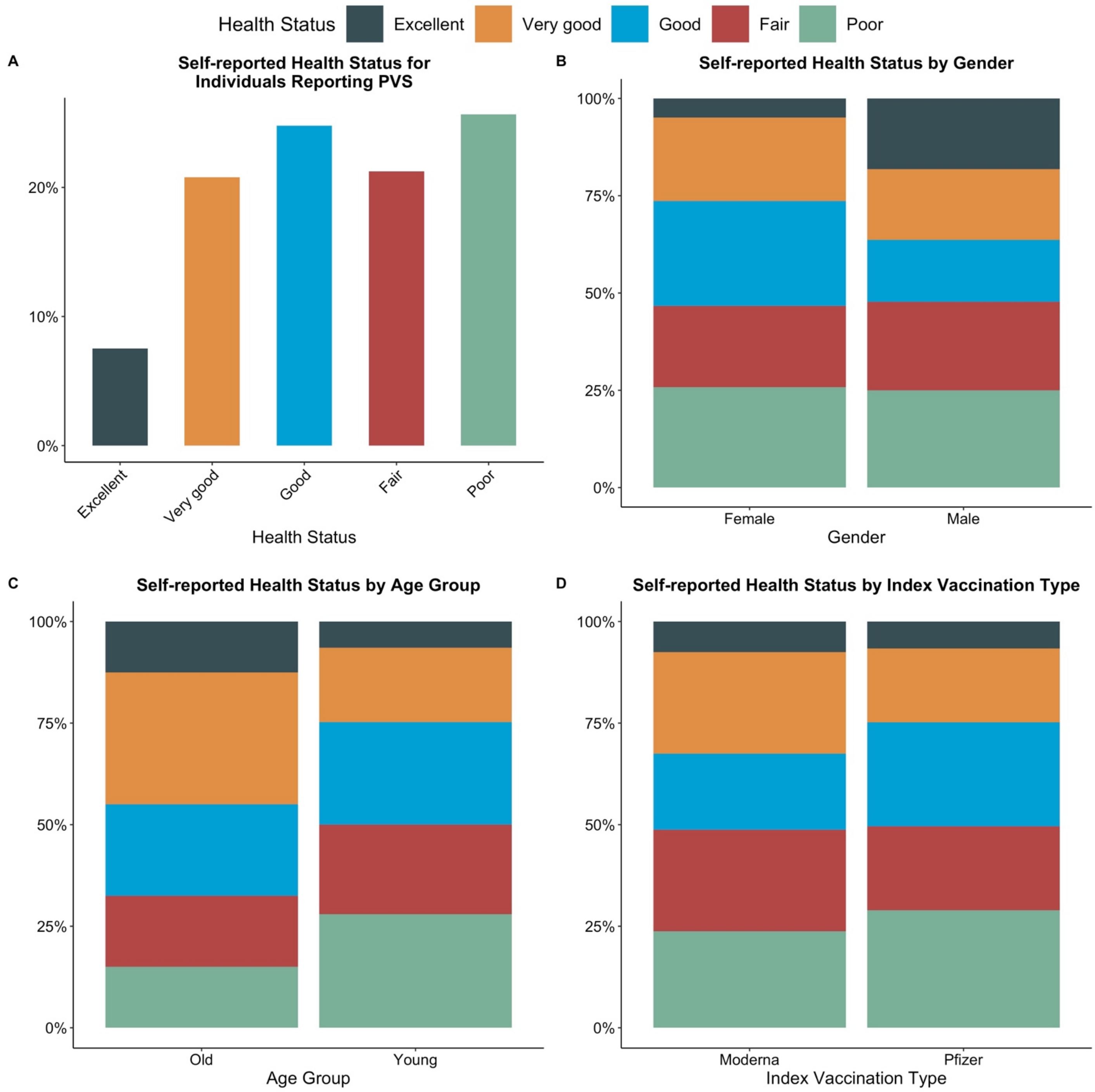

For (**A**) all participants reporting PVS, (**B**) all participants reporting PVS stratified by gender, (**C**) all participants reporting PVS stratified by age group, and (**D**) all participants reporting PVS stratified by index vaccination type. Assessed by the question, "Would you say your health in general is…" followed by six options: excellent, very good, good, fair, poor, or don't know. The 15 respondents answering "don't know" are excluded from the calculations used to produce these figures.
PVS, Post-vaccination Syndrome

**Supplemental Figure 5. Distribution of symptom severity**.

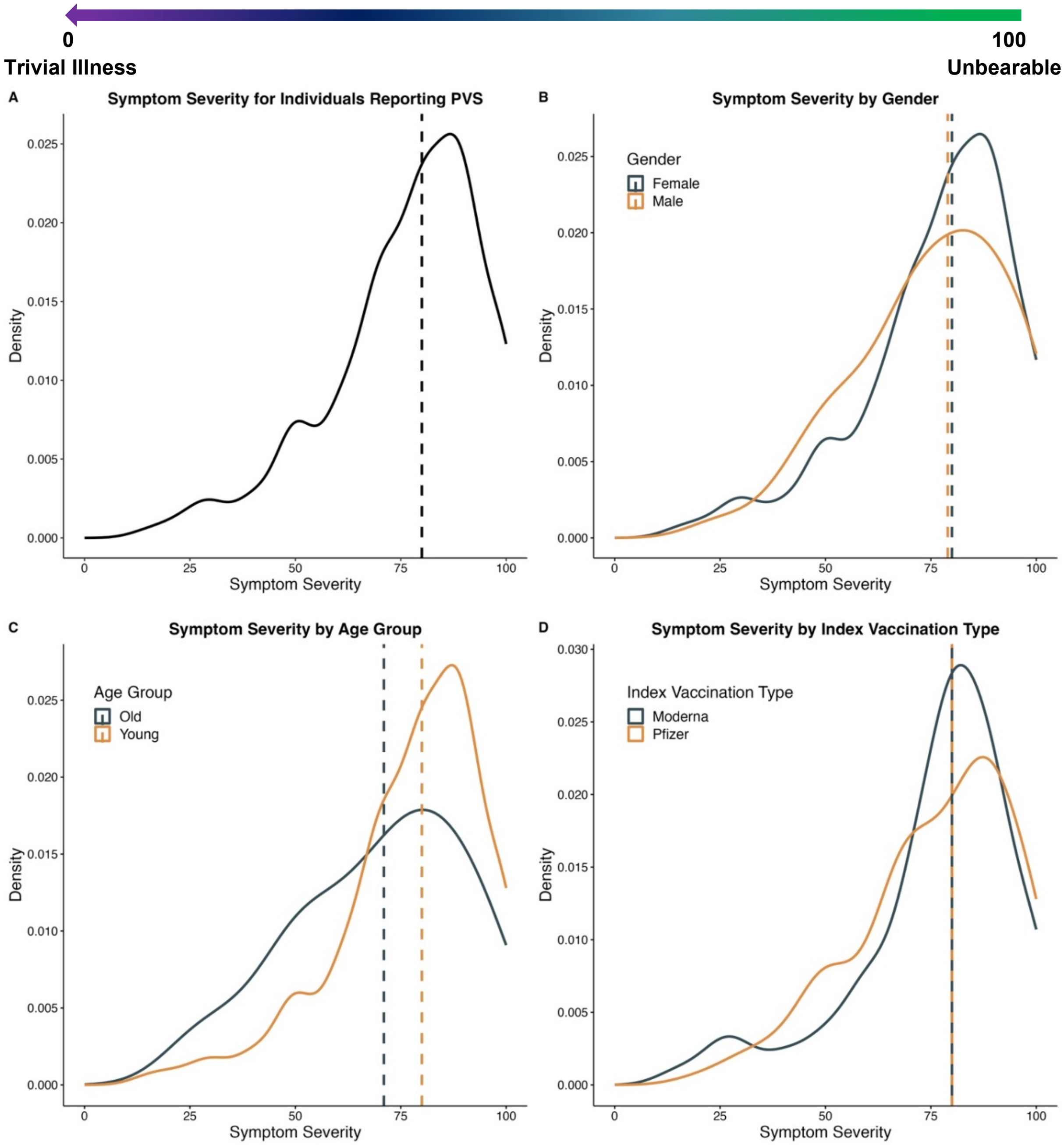

For (**A**) all participants who reported PVS, (**B**) all participants who reported PVS stratified by gender, (**C**) all participants who reported PVS stratified by age group, and (**D**) all participants who reported PVS stratified by index vaccination type. Median values are indicated with dashed lines. Assessed by the question, "We are trying to get a sense of how bad your PVS symptoms are when you feel them the most. On the slider below, with 0 being a trivial illness and 100 being unbearable, please let us know what the worst days are like." (**Supplemental Appendix** [Health status questions]). PVS, Post-vaccination Syndrome

**Supplemental Figure 6. Symptom prevalence among 241 participants who reported PVS.**

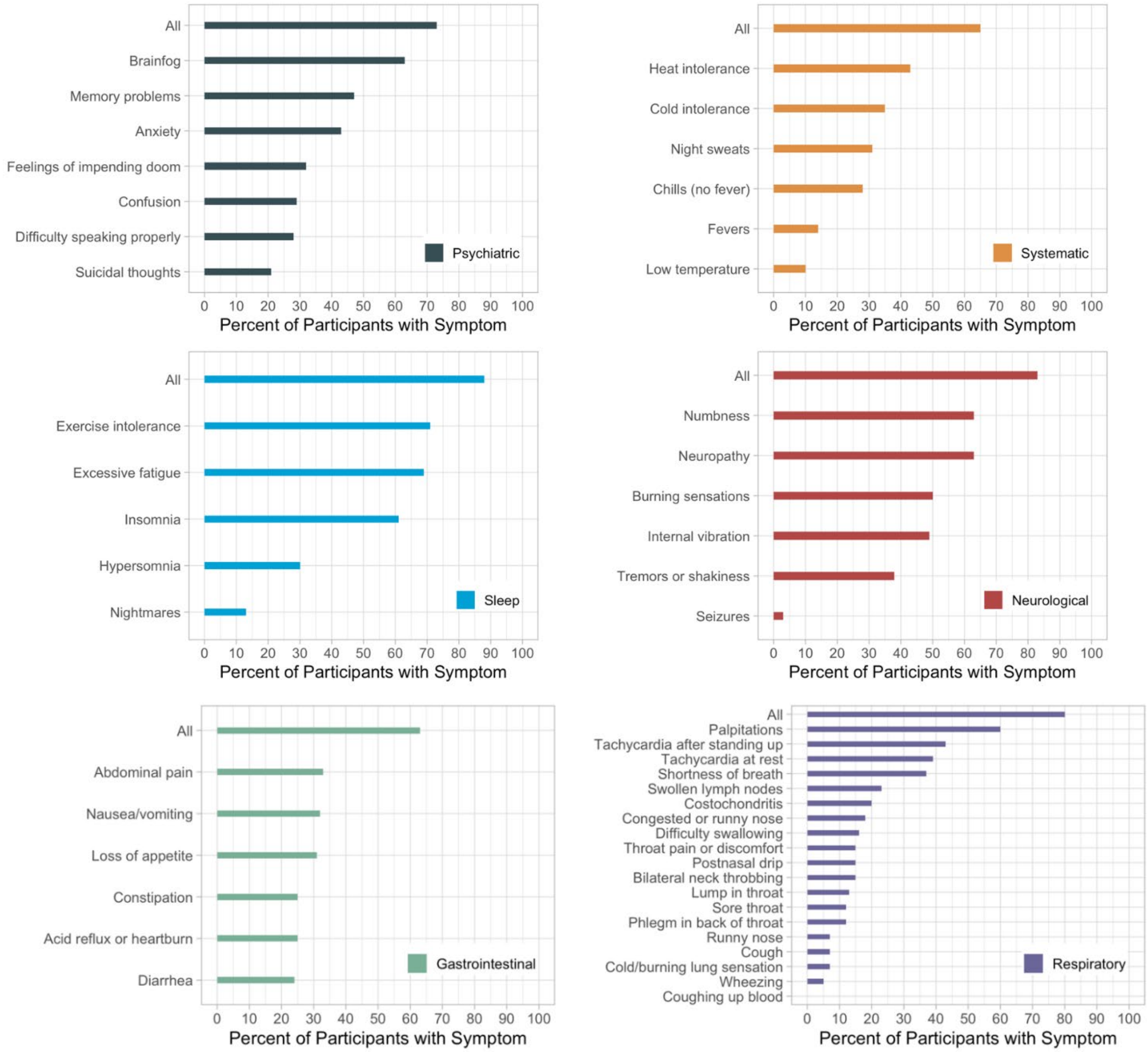

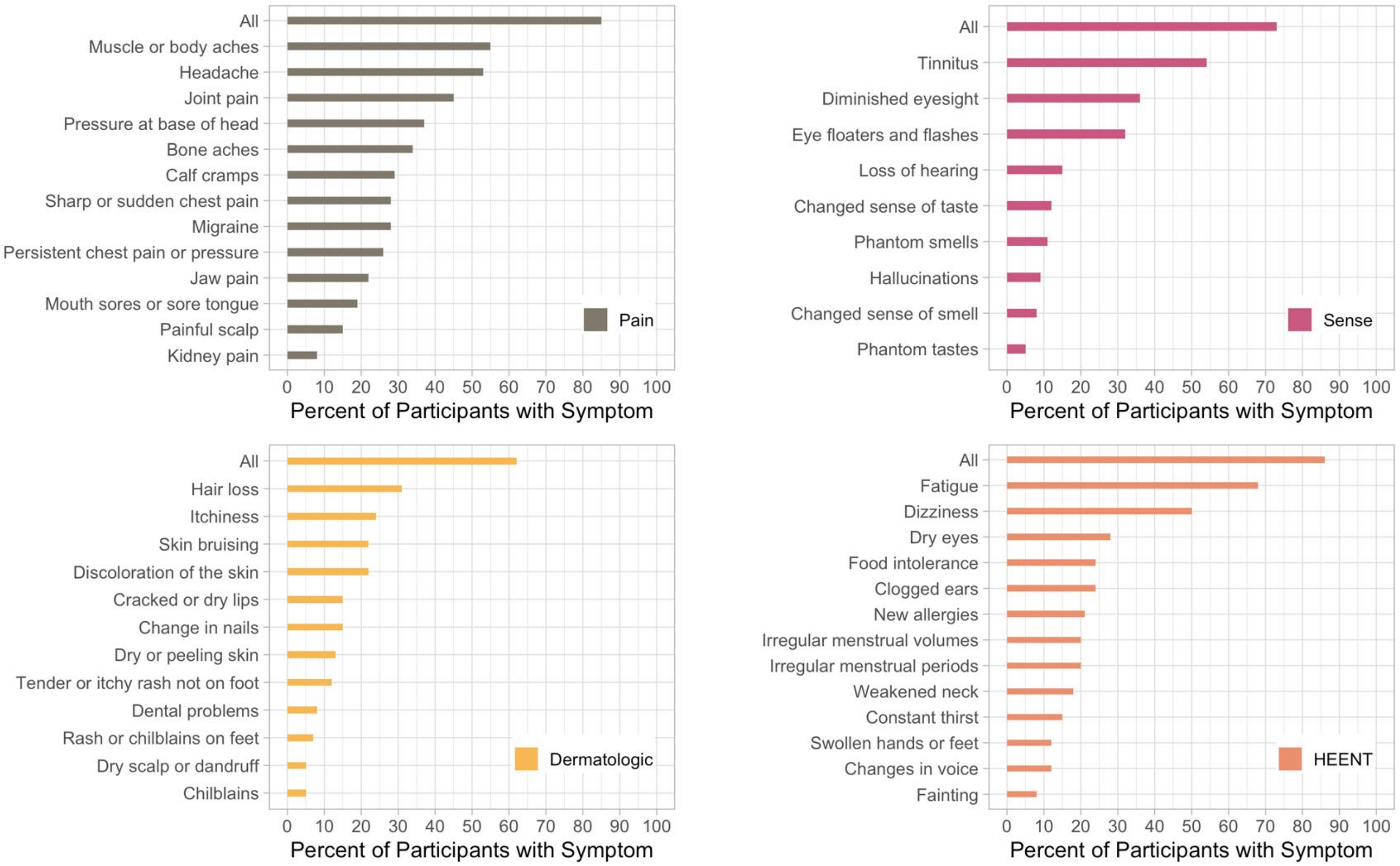

"All" row means participants who reported any of the symptoms in the corresponding symptom group. HEENT, Head, Eyes, Ears, Nose, Throat; PVS, Post-vaccination Syndrome

**Supplemental Table 1. Participant pre-pandemic comorbidity frequencies (N = 240).***

| Comorbidity | n | % |
|---|---|---|
| Any allergies | 116 | 48 |
| Arthritis (including rheumatoid arthritis, gout, lupus, or fibromyalgia) | 31 | 13 |
| Asthma | 49 | 20 |
| Autoimmune disease (including lupus, scleroderma, etc.) | 26 | 11 |
| Bleeding disorder (including sickle cell disease or Thalassemia) | 3 | 1 |
| Blood clots | 2 | 1 |
| Cancer or malignancy of any kind | 13 | 5 |
| Cerebrovascular conditions affecting blood vessels to or in the brain (including stroke) | 2 | 1 |
| Chronic lung disease (including emphysema, chronic bronchitis, COPD, or pulmonary fibrosis) | 3 | 1 |
| Cystic fibrosis | 0 | 0 |
| Diabetes | 6 | 3 |
| Ehlers Danlos Syndrome (hypermobile joints) | 8 | 3 |
| Gastrointestinal issues (including IBS or acid reflux) | 67 | 28 |
| Heart attack, also called myocardial infarction | 2 | 1 |
| Heart conditions (including coronary artery disease or cardiomyopathies) | 8 | 3 |
| Heart failure | 0 | 0 |
| High cholesterol | 34 | 14 |
| History of organ transplant (including kidney, liver, heart, or lung) | 0 | 0 |
| Hypertension or high blood pressure | 31 | 13 |
| Immunocompromised state (including weakened immune system from blood or bone marrow transplant, immune deficiencies, HIV, use of corticosteroids, or use of other immune-weakening medicines) | 6 | 3 |
| Kidney disease | 1 | 0.4 |
| Liver disease | 0 | 0 |
| Lyme disease | 7 | 3 |
| MCAS or other mast cell disorders | 3 | 1 |
| ME/CFS | 9 | 4 |

| | | |
|---|---|---|
| Migraines | 46 | 19 |
| Neurologic conditions (including seizures, dementia, multiple sclerosis, Parkinson's, neuropathy, small fiber neuropathy, etc.) | 9 | 4 |
| POTS or dysautonomia | 8 | 3 |
| Spinal disorder(s) | 9 | 4 |
| Tremors/Internal vibrations | 6 | 3 |
| Depressive disorders | 49 | 20 |
| Anxiety disorders | 61 | 25 |
| Schizophrenia spectrum and other psychotic disorders | 0 | 0 |
| Bipolar and related disorders | 1 | 0.4 |
| Obsessive-Compulsive and related disorders | 6 | 3 |
| Trauma- and stressor-related disorders | 17 | 7 |
| Feeding and eating disorders | 5 | 2 |
| Somatic symptoms (excessive thoughts, feelings and behaviors relating to the physical symptoms) and related disorders | 2 | 1 |

*One participant did not have a response for the pre-pandemic comorbidities. After excluding any allergies, high cholesterol, and hypertension, 59 out of 240 (25%) participants were found without any listed comorbidities.

COPD, chronic obstructive pulmonary disease; HIV, human immunodeficiency virus; IBS, irritable bowel syndrome; MCAS, mast cell activation syndrome; ME/CFS, myalgic encephalomyelitis/chronic fatigue syndrome; POTS, postural orthostatic hypotension

**Supplemental Table 2. Participant experience (N = 241).**

| Characteristic | n | % |
|---|---|---|
| **Felt fearful** | | |
| Always | 14 | 6 |
| Often | 46 | 19 |
| Sometimes | 89 | 37 |
| Rarely | 45 | 19 |
| Never | 44 | 18 |
| Missing | 3 | – |
| **Felt anxious** | | |
| Always | 8 | 3 |
| Often | 24 | 10 |
| Sometimes | 74 | 31 |
| Rarely | 74 | 31 |
| Never | 58 | 24 |
| Missing | 3 | – |
| **Felt worried** | | |
| Always | 9 | 4 |
| Often | 38 | 16 |
| Sometimes | 77 | 32 |
| Rarely | 68 | 29 |
| Never | 46 | 19 |
| Missing | 3 | – |
| **Felt unease** | | |
| Always | 14 | 6 |
| Often | 62 | 26 |
| Sometimes | 102 | 43 |
| Rarely | 43 | 18 |
| Never | 17 | 7 |
| Missing | 3 | – |
| **Felt worthless** | | |
| Always | 4 | 2 |
| Often | 17 | 7 |

| | | |
|---|---|---|
| Sometimes | 48 | 20 |
| Rarely | 47 | 20 |
| Never | 122 | 51 |
| Missing | 3 | – |
| **Felt helpless** | | |
| Always | 10 | 4 |
| Often | 63 | 26 |
| Sometimes | 80 | 34 |
| Rarely | 37 | 16 |
| Never | 48 | 20 |
| Missing | 3 | – |
| **Felt depressed** | | |
| Always | 12 | 5 |
| Often | 34 | 14 |
| Sometimes | 82 | 34 |
| Rarely | 54 | 23 |
| Never | 56 | 24 |
| Missing | 3 | – |
| **Felt hopeless** | | |
| Always | 6 | 3 |
| Often | 44 | 18 |
| Sometimes | 67 | 28 |
| Rarely | 54 | 23 |
| Never | 67 | 28 |
| Missing | 3 | – |
| **Felt fatigued** | | |
| Always | 63 | 26 |
| Often | 93 | 39 |
| Sometimes | 58 | 24 |
| Rarely | 21 | 9 |
| Never | 3 | 1 |
| Missing | 3 | – |
| **Felt tired** | | |
| Always | 32 | 13 |

| | | |
|---|---|---|
| Often | 94 | 39 |
| Sometimes | 69 | 29 |
| Rarely | 33 | 14 |
| Never | 10 | 4 |
| Missing | 3 | – |
| **Felt rundown** | | |
| Very much | 55 | 23 |
| Quite a bit | 90 | 38 |
| Somewhat | 49 | 21 |
| A little bit | 39 | 16 |
| Not at all | 5 | 2 |
| Missing | 3 | – |
| **Had difficulty falling asleep** | | |
| Very much | 43 | 18 |
| Quite a bit | 46 | 19 |
| Somewhat | 64 | 27 |
| A little bit | 63 | 26 |
| Not at all | 22 | 9 |
| Missing | 3 | – |
| **Pain interfered with activities** | | |
| Very much | 34 | 14 |
| Quite a bit | 58 | 24 |
| Somewhat | 62 | 26 |
| A little bit | 50 | 21 |
| Not at all | 34 | 14 |
| Missing | 3 | – |
| **Number of close people to rely on for help** | | |
| 1-2 | 98 | 41 |
| 3-5 | 98 | 41 |
| 5+ | 39 | 16 |
| None | 3 | 1 |
| Missing | 3 | – |
| **Had neighbors willing to help** | | |
| Difficult | 46 | 19 |

| | | |
|---|---|---|
| Easy | 32 | 13 |
| Possible | 106 | 45 |
| Very difficult | 40 | 17 |
| Very easy | 14 | 6 |
| Missing | 3 | – |
| **Had someone to help** | | |
| Always | 78 | 33 |
| Usually | 92 | 39 |
| Sometimes | 44 | 18 |
| Rarely | 15 | 6 |
| Never | 9 | 4 |
| Missing | 3 | – |
| **Lacked companionship** | | |
| Hardly ever or never | 103 | 43 |
| Often | 47 | 20 |
| Some of the time | 88 | 37 |
| Missing | 3 | – |
| **Felt left out** | | |
| Hardly ever or never | 70 | 30 |
| Often | 55 | 23 |
| Some of the time | 112 | 47 |
| Missing | 4 | – |
| **Felt isolated** | | |
| Hardly ever or never | 52 | 22 |
| Often | 77 | 32 |
| Some of the time | 109 | 46 |
| Missing | 3 | – |
| **Felt lonely** | | |
| Hardly ever | 53 | 22 |
| Never | 27 | 11 |
| Occasionally | 77 | 32 |
| Often/Always | 28 | 12 |
| Some of the time | 53 | 22 |
| Missing | 3 | – |

| | | |
|---|---|---|
| **Living situation** | | |
| I do not have a steady place to live | 1 | 0.4 |
| I have a place to live today, but I am worried about losing it in the future | 16 | 7 |
| I have a steady place to live | 221 | 93 |
| Missing | 3 | – |
| **Food insecurity** | | |
| Never true | 217 | 91 |
| Often true | 4 | 2 |
| Sometimes true | 17 | 7 |
| Missing | 3 | – |
| **Unmet medical transport** | | |
| No | 231 | 96 |
| Yes | 10 | 4 |
| **Unmet non-medical transport** | | |
| No | 228 | 95 |
| Yes | 13 | 5 |

**Supplemental Table 3. Health status.**

| Characteristic | n | % |
|---|---|---|
| **Euro-QoL visual analogue scale (0-100)**[1, 2] | 50 (39-70) | – |
| **How bad are your PVS or other symptoms (0-100) on your worst days?**[1, 3] | 80 (69-89) | – |
| Missing | 3 | – |
| **Self-reported health status**[1] | | |
| Don't know | 15 | 6 |
| Excellent | 17 | 7 |
| Fair | 48 | 20 |
| Good | 56 | 23 |
| Poor | 58 | 24 |
| Very good | 47 | 20 |

[1]Median (IQR) EQ-VAS and symptom severity; n/N, % for self-reported health status

[2]The Euro-QoL visual analogue scale is measured from 0 (worst imaginable health) to 100 (best imaginable health).

[3]Symptom severity is measured from 0 (trivial illness) to 100 (unbearable).

PVS, Post-vaccination Syndrome

**Supplemental Table 4. Symptom frequency of PVS.**

| Symptom | n | % |
|---|---|---|
| Abdominal pain | 80 | 33 |
| Acid reflux or heartburn | 61 | 25 |
| Anxiety | 104 | 43 |
| Bilateral neck throbbing around lymph nodes | 35 | 15 |
| Bone aches | 81 | 34 |
| Brain fog | 151 | 63 |
| Burning sensations | 121 | 50 |
| Calf cramps | 71 | 29 |
| Change in nails | 37 | 15 |
| Changed sense of smell | 20 | 8 |
| Changed sense of taste | 28 | 12 |
| Changes in voice | 30 | 12 |
| Chilblains | 11 | 5 |
| Chills (no fever) | 68 | 28 |
| Clogged ears | 59 | 24 |
| Cold intolerance | 85 | 35 |
| Cold or burning feeling in lungs | 16 | 7 |
| Confusion | 71 | 29 |
| Congested or runny nose | 44 | 18 |
| Constant thirst | 37 | 15 |
| Constipation | 60 | 25 |
| Costochondritis | 49 | 20 |
| Cough | 18 | 7 |
| Coughing up blood | 1 | 0.4 |
| Cracked or dry lips | 35 | 15 |
| Dental problems | 20 | 8 |
| Diarrhea | 59 | 24 |
| Difficulty speaking properly | 67 | 28 |
| Difficulty swallowing | 38 | 16 |
| Discoloration of the skin | 52 | 22 |
| Dizziness | 121 | 50 |
| Dry eyes | 67 | 28 |
| Dry or peeling skin | 32 | 13 |
| Dry scalp or dandruff | 11 | 5 |
| Excessive fatigue | 167 | 69 |
| Exercise intolerance | 170 | 71 |
| Fainting | 20 | 8 |

| | | |
|---|---|---|
| Fatigue | 165 | 68 |
| Feelings of impending doom | 77 | 32 |
| Fevers | 33 | 14 |
| Floaters or flashes of light in vision | 77 | 32 |
| Hair loss | 74 | 31 |
| Hallucinations | 21 | 9 |
| Headache | 128 | 53 |
| Heat intolerance | 104 | 43 |
| Hypersomnia | 72 | 30 |
| Inability to eat or tolerate food | 58 | 24 |
| Insomnia | 148 | 61 |
| Internal tremors or buzzing/vibration | 118 | 49 |
| Irregular or skipped menstrual cycles | 49 | 20 |
| Itchiness | 57 | 24 |
| Jaw pain | 53 | 22 |
| Joint pain | 108 | 45 |
| Kidney pain | 20 | 8 |
| Loss of appetite | 74 | 31 |
| Loss of hearing | 35 | 15 |
| Loss or decrease in quality of vision | 86 | 36 |
| Low temperature | 23 | 10 |
| Lump in throat | 31 | 13 |
| Memory problems | 113 | 47 |
| Menstrual cycles that are heavier or lighter than normal | 49 | 20 |
| Migraine | 68 | 28 |
| Mouth sores or sore tongue | 46 | 19 |
| Muscle or body aches | 132 | 55 |
| Nausea/vomiting | 76 | 32 |
| Neuropathy | 151 | 63 |
| New allergies | 51 | 21 |
| Night sweats | 74 | 31 |
| Nightmares | 32 | 13 |
| Numbness | 153 | 63 |
| Painful scalp | 37 | 15 |
| Palpitations | 145 | 60 |
| Persistent chest pain or pressure | 63 | 26 |
| Phantom smells | 26 | 11 |
| Phantom tastes | 11 | 5 |
| Phlegm in back of throat | 30 | 12 |
| Postnasal drip | 35 | 15 |
| Pressure at base of head | 90 | 37 |

| | | |
|---|---|---|
| Runny nose | 17 | 7 |
| Seizures | 7 | 3 |
| Sharp or sudden chest pain | 68 | 28 |
| Shortness of breath or difficulty breathing | 88 | 37 |
| Skin bruising | 53 | 22 |
| Sore throat | 28 | 12 |
| Suicidal thoughts | 50 | 21 |
| Swollen hands or feet | 30 | 12 |
| Swollen lymph nodes | 55 | 23 |
| Tachycardia after standing up | 103 | 43 |
| Tachycardia at rest | 94 | 39 |
| Tender or itchy rash not on foot | 28 | 12 |
| Tender or itchy rash or chilblains on the toes or foot | 16 | 7 |
| Throat pain or discomfort | 36 | 15 |
| Tinnitus or humming in ears | 131 | 54 |
| Tremors or shakiness | 91 | 38 |
| Weakened neck | 43 | 18 |
| Wheezing | 13 | 5 |

PVS, Post-vaccination Syndrome

**Supplemental Table 5. Participant new-onset conditions (N = 240).**[1]

| New-onset Condition | n (%) |
|---|---|
| Any allergies | 108 (45) |
| Arthritis (including rheumatoid arthritis, gout, lupus, or fibromyalgia) | 42 (17) |
| Asthma | 37 (15) |
| Autoimmune disease (including lupus, scleroderma, etc.) | 47 (20) |
| Bleeding disorder (including sickle cell disease or Thalassemia) | 2 (1) |
| Blood clots | 10 (4) |
| Cancer or malignancy of any kind | 7 (3) |
| Cerebrovascular conditions affecting blood vessels to or in the brain (including stroke) | 13 (5) |
| Chronic lung disease (including emphysema, chronic bronchitis, COPD, or pulmonary fibrosis) | 5 (2) |
| Cystic fibrosis | 0 (0) |
| Diabetes | 6 (3) |
| Ehlers Danlos Syndrome (hypermobile joints) | 15 (6) |
| Gastrointestinal issues (including IBS or acid reflux) | 73 (30) |
| Heart attack/myocardial infarction | 3 (1) |
| Heart conditions (including coronary artery disease or cardiomyopathies) | 31 (13) |
| Heart failure | 3 (1) |
| High cholesterol | 52 (22) |
| History of organ transplant (including kidney, liver, heart, or lung) | 0 (0) |
| Hypertension or high blood pressure | 45 (19) |
| Immunocompromised state (including weakened immune system from blood or bone marrow transplant, immune deficiencies, HIV, use of corticosteroids, or use of other immune-weakening medicines) | 22 (9) |
| Kidney disease | 2 (1) |
| Liver disease | 6 (3) |
| Long COVID | 21 (9) |
| Lyme disease | 9 (4) |
| MCAS or other mast cell disorders | 28 (12) |
| ME/CFS | 28 (12) |
| Migraines | 53 (22) |
| Neurological conditions (including seizures, dementia, multiple sclerosis, Parkinson's, neuropathy, small fiber neuropathy, etc.) | 79 (33) |
| POTS or dysautonomia | 70 (29) |
| Spinal disorder(s) | 13 (5) |
| Vaccine injury | 133 (55) |
| Depressive disorders | 49 (20) |
| Anxiety disorders | 86 (36) |
| Schizophrenia spectrum and other psychotic disorders | 0 (0) |

| | |
|---|---|
| Bipolar and related disorders | 1 (0.4) |
| Obsessive-Compulsive and related disorders | 6 (3) |
| Trauma- and stressor-related disorders | 25 (10) |
| Feeding and eating disorders | 4 (2) |
| Somatic symptom and related disorders | 4 (2) |

[1]One participant did not provide a response for new onset medical conditions.
COPD, chronic obstructive pulmonary disease; HIV, human immunodeficiency virus; IBS, irritable bowel syndrome; MCAS, mast cell activation syndrome; ME/CFS, myalgic encephalomyelitis/chronic fatigue syndrome; POTS, postural orthostatic hypotension

**Supplemental Table 6. Frequency of treatments tried by participants with PVS.**

| Category | Treatment | n (%) |
|---|---|---|
| Vitamins and Supplements | Vitamin B1 (Thiamine) | 52 (22) |
| | Vitamin B2 (Riboflavin) | 42 (17) |
| | Vitamin B3 (Niacin) | 36 (15) |
| | Vitamin B6 (Pyridoxine) | 39 (16) |
| | Vitamin B complex | 102 (42) |
| | Vitamin B12 (hydroxocobalamin, cyanocobalamin) | 103 (43) |
| | Vitamin C (ascorbic acid) | 128 (53) |
| | Vitamin D (calciferol) | 157 (65) |
| | Vitamin E (alpha-tocopherol) | 22 (9) |
| | Vitamin K2 (menaquinone) | 52 (22) |
| | Zinc | 86 (36) |
| | Copper | 12 (5) |
| | CoQ10 | 74 (31) |
| | Magnesium | 135 (56) |
| | Alpha lipoic acid | 62 (26) |
| | Flavonoids (Quercetin, Luteolin, BrainGain) | 71 (29) |
| | NAC (N-Acetyl Cysteine)/Glycine | 79 (33) |
| | Glutathione | 47 (20) |
| | Glutamine | 10 (4) |
| | PEA (Palmitoylethanolamide) | 17 (7) |
| | Turmeric (Curcumin) | 96 (40) |
| | Omega 3s/Fish oil | 97 (40) |
| | Acetyl-L-Carnitine | 30 (12) |
| | L-Lysine | 25 (10) |
| H1 Antihistamines | Loratadine (Claritin) | 84 (35) |
| | Cetirizine (Zyrtec) | 87 (36) |
| | Fexofenadine (Allegra) | 39 (16) |
| | Diphenhydramine (Benadryl) | 67 (28) |
| | Chlorpheniramine (Chlor-Trimeton) | 3 (1) |
| | Hydroxyzine (Atarax) (Rx) | 22 (9) |
| H2 Antihistamines | Famotidine (Pepcid) | 91 (38) |

| | | |
|---|---|---|
| | Cimetidine (Tagamet) | 4 (2) |
| Motion Sickness Drugs | Dimenhydrinate (Dramamine) | 11 (5) |
| | Meclizine (Bonine) | 17 (7) |
| | Scopolamine | 2 (1) |
| NSAIDs | Ibuprofen (Advil, Motrin, Provil) | 129 (54) |
| | Naproxen (Aleve, Mediproxen, All Day Pain Relief) | 58 (24) |
| | Aspirin [Acetylsalicylic acid] (Ascriptin, Aspercin, Aspirtab, Buffasal, Bufferin, Buffinol, Easprin, Ecotrin, Vazalore), 81 mg (baby aspirin), 325 mg (regular aspirin) | 84 (35) |
| Non-NSAID Painkillers | Acetaminophen [Paracetamol] (Tylenol, Panadol, Apra, FeverAll, Mapap, Pharbetol) | 111 (46) |
| Herbal and Natural Supplements | Black seed oil | 43 (18) |
| | Milk thistle | 27 (11) |
| | Astaxanthin | 7 (3) |
| | Nattokinase | 32 (13) |
| | Moringa | 9 (4) |
| | Melatonin | 79 (33) |
| | Reishi, Lion’s Mane, Cordyceps (mushrooms) | 28 (12) |
| | CBD (oral, topical, inhaled) | 68 (28) |
| | Cannabis | 33 (14) |
| | Eastern white pine needle | 19 (8) |
| | Nettles | 13 (5) |
| | Dandelion | 32 (13) |
| | St. John’s Wort (hypericum perforatum) | 4 (2) |
| Probiotics | | 238 (99) |
| Mast Cell Stabilizers | Cromoglicic acid (Cromolyn) oral, nasal spray | 20 (8) |
| | Ketotifen (Zaditor) | 12 (5) |
| | Montelukast (Singulair) | 22 (9) |
| | Omalizumab (Xolair) | 1 (0.4) |
| | DAO enzyme supplement (OTC) | 19 (8) |
| Antibiotics | Macrolides (Azithromycin, Clarithromycin, Erythromycin) | 7 (3) |
| | Fluoroquinolones (Ciprofloxacin, Levofloxacin, other) | 2 (1) |
| | Cephalosporins (Cephalexin, Ceftriaxone, other) | 2 (1) |
| | Penicillin (Penicillin, Amoxicillin, other) | 10 (4) |
| | Doxycycline | 17 (7) |

| | | |
|---|---|---|
| Antiparasitic and Antimalarial Treatments | Ivermectin (Stromectol) | 44 (18) |
| | Hydroxychloroquine (Plaquenil) | 19 (8) |
| | Chloroquine (Aralen) | 0 (0) |
| Steroids | Oral (Prednisone, Dexamethasone, Hydrocortisone, Methylprednisolone, other) | 116 (48) |
| | IV steroids (Hydrocortisone, Methylprednisolone, other) | 12 (5) |
| | Inhaled steroids (Mometasone [Asmanex], Fluticasone [Flovent], Budesonide [Pulmicort], other) | 24 (10) |
| | Other steroid injections (intratympanic, spinal) | 9 (4) |
| Anticoagulants | Heparin | 3 (1) |
| | Warfarin (Coumadin) | 0 (0) |
| | Direct oral anticoagulant: Apixaban (Eliquis) | 2 (1) |
| | Direct oral anticoagulant: Dabigatran (Pradaxa) | 1 (0.4) |
| | Direct oral anticoagulant: Rivaroxaban (Xarelto) | 4 (2) |
| | Direct oral anticoagulant: Edoxaban (Savaysa) | 0 (0) |
| | Direct oral anticoagulant: Betrixaban (Bevyxxa) | 0 (0) |
| Anti-Platelet Therapies | Clopidogrel (Plavix) | 7 (3) |
| Anti-Inflammatory Drugs | Colchicine (Colcrys, Mitigare, Gloperba) | 14 (6) |
| Antiviral Drugs for COVID-19 | Nirmatrelvir/Ritonavir (Paxlovid) | 4 (2) |
| | Molnupiravir (Lagevrio) | 0 (0) |
| | Remdesivir (Veklury) | 0 (0) |
| Antivirals for Herpesvirus | Valacyclovir (Valtrex) | 23 (10) |
| | Famciclovir (Famvir) | 3 (1) |
| | Acyclovir (Zovirax) | 10 (4) |
| Monoclonal Antibody Therapies | Casirivimab/Imdevimab (Regeneron) | 7 (3) |
| | Bamlanivimab and etesevimab | 2 (1) |
| | Sotrovimab (Xevudy) | 0 (0) |
| | Bebtelovimab | 0 (0) |
| | Tixagevimab and cilgavimab (Evusheld) | 4 (2) |
| Blood Derived Treatments | Intravenous Immunoglobulin (IVIG) | 17 (7) |
| | Subcutaneous Immunoglobulin (SCIG) | 2 (1) |
| | Convalescent plasma | 0 (0) |
| Immunomodulatory Drugs | Eculizumab (Soliris) | 0 (0) |
| | IL-1 antagonist (Anakinra [Kineret], Canakinumab [Ilaris], other) | 2 (1) |

| | | |
|---|---|---|
| | IL-6 antagonist (Siltuximab [Sylvant], Sarilumab [Kevzara], Tocilizumab [Actemra], other) | 0 (0) |
| | Kinase inhibitors (Acalabrutinib [Calquence], Ibrutinib [Imbruvica], Zanubrutinib [Brukinsa], Baricitinib [Olumiant], Ruxolitinib [Jakafi], Tofacitinib [Xeljanz], other) | 1 (0.4) |
| | T-Cell and B-Cell inhibitors (mycophenolate mofetil [CellCept]) | 2 (1) |
| | Monoclonal immunosuppressives: CD-20/B-Cell modulator | 1 (0.4) |
| | Monoclonal immunosuppressives: Rituximab (Riabni, Rituxan, Ruxience, Truxima) | 0 (0) |
| | Monoclonal immunosuppressives: Leronlimab | 0 (0) |
| | CCR5 entry inhibitors: Maraviroc (Selzentry) | 11 (5) |
| G-Protein Coupled Receptor Medications | BC 007 | 0 (0) |
| Anticonvulsant | Gabapentin (Neurontin, Gralise) | 61 (25) |
| | Pregabalin (Lyrica) | 13 (5) |
| | Lamotrigine (Lamictal) | 2 (1) |
| | Carbamazepine (Tegretol) | 2 (1) |
| Antidepressants | Duloxetine (Cymbalta) | 22 (9) |
| | Doxepin (Sinequan) | 2 (1) |
| | Amitriptyline (Elavil) | 15 (6) |
| | Nortriptyline (Pamelor) | 7 (3) |
| | Fluvoxamine (Luvox) | 16 (7) |
| | Mirtazapine (Remeron) | 2 (1) |
| | Fluoxetine (Prozac) | 8 (3) |
| | Escitalopram (Lexapro) | 18 (7) |
| | Citalopram (Celexa) | 5 (2) |
| | Paroxetine (Brisdelle, Paxil, Pexeva) | 1 (0.4) |
| | Sertraline (Zoloft) | 11 (5) |
| | Desvenlafaxine (Pristiq) | 1 (0.4) |
| | Venlafaxine (Effexor) | 8 (3) |
| Benzodiazepines | Alprazolam (Xanax) | 15 (6) |
| | Clonazepam (Klonopin) | 19 (8) |
| | Lorazepam (Ativan) | 21 (9) |
| | Diazepam (Valium) | 6 (2) |
| | Triazolam (Halcion) | 0 (0) |

| | | |
|---|---|---|
| | Temazepam (Restoril) | 1 (0.4) |
| Muscle Relaxants | Baclofen (Lioresal) | 8 (3) |
| | Methocarbamol (Robaxin) | 5 (2) |
| | Cyclobenzaprine (Flexeril) | 22 (9) |
| | Metaxalone (Skelaxin) | 1 (0.4) |
| | Tizanidine (Zanaflex) | 13 (5) |
| Sleep Medications | Zolpidem (Ambien) | 13 (5) |
| | Zaleplon (Sonata) | 0 (0) |
| | Trazodone (Desyrel) | 12 (5) |
| Opioid Antagonists | Low dose Naltrexone | 48 (20) |
| Beta Blockers | Propranolol (Inderal) | 27 (11) |
| | Metoprolol (Lopressor) | 24 (10) |
| | Nebivolol (Bystolic) | 2 (1) |
| Other Blood Pressure Medications | Lisinopril (Prinivil, Zestril) | 7 (3) |
| | Enalapril (Epaned) | 0 (0) |
| | Candesartan (Atacand) | 4 (2) |
| | Losartan (Cozaar) | 4 (2) |
| | Amlodipine (Norvasc) | 5 (2) |
| | Verapamil (Calan, Isoptin) | 4 (2) |
| | Diltiazem (Cardizem) | 5 (2) |
| Diuretics | Furosemide (Lasix) | 2 (1) |
| | Bumetanide (Bumex) | 0 (0) |
| | Hydrochlorothiazide (Microzide) | 3 (1) |
| | Chlorthalidone (Thalitone) | 0 (0) |
| | Acetazolamide (Diamox) | 3 (1) |
| | Spironolactone (Aldactone, CaroSpir) | 7 (3) |
| Salt Supplements | SaltStick | 11 (5) |
| Anti-Hypotensive Treatments | Midodrine (Proamatine) | 7 (3) |
| Antiarrhythmics | Amiodarone (Nexterone) | 0 (0) |
| | Flecainide (Tambocor) | 2 (1) |
| Lipid Lowering Medications | Pravastatin (Pravachol) | 11 (5) |
| | Atorvastatin (Lipitor) | 17 (7) |
| | Rosuvastatin (Crestor) | 7 (3) |

| | | |
|---|---|---|
| Other Respiratory Treatments | Bronchodilators (Albuterol, Salmeterol, Formoterol, Ipratropium, Tiotropium, other) | 26 (11) |
| | Nasal spray (Flonase [Fluticasone], Nasocort [Triamcinolone], Afrin [Oxymetazoline], other) | 59 (24) |
| Other Gastrointestinal Treatments | Proton pump inhibitors (Omeprazole [Prilosec], Esomeprazole [Nexium]) | 43 (18) |
| | Anti-nausea medication (Ondansetron [Zofran], Prochlorperazine [Compazine], Promethazine [Phenergan], Metoclopramide [Reglan]) | 25 (10) |
| | Antispasmodics (Hyoscyamine [Levsin], Dicyclomine [Bentyl], Clidinium and chlordiazepoxide [Librax], atropine, scopolamine, and phenobarbital [Donnatal]) | 7 (3) |
| | Laxatives, stool softeners | 32 (13) |
| | Antidiarrheal (Diphenoxylate [Lomotil], Loperamide [Imodium], Bismuth subsalicylate [Kaopectate, Pepto Bismol]) | 10 (4) |
| | Treatment for SIBO (Rifaximin [Xifaxan], other) | 4 (2) |
| | Simethicone (Gas-X) | 16 (7) |
| Psychedelics | Lysergic Acid Diethylamide (LSD) | 0 (0) |
| | Psilocin (4-HO-DMT) | 3 (1) |
| | 3, 4-Methylenedioxymethamphetamine (MDMA) | 0 (0) |
| | Ayahuasca | 0 (0) |
| Pain Medications | Lidocaine | 16 (7) |
| | Capsaicin | 8 (3) |
| | Tramadol (ConZip, Qdolo, Ultram) | 4 (2) |
| | Ketorolac (Toradol) | 6 (2) |
| | Celecoxib (Celebrex, Elyxyb) | 8 (3) |
| | Opioid analgesics (Morphine, Buprenorphine, Codeine, Fentanyl, Oxycodone, Hydrocodone, Hydromorphone, other) | 10 (4) |
| Other Medications | Ketamine (Ketalar) | 2 (1) |
| | Botulinum toxin injection (Botox) | 9 (4) |
| Non-Pharmacological Agents | Enhanced External Counterpulsation (EECP) | 1 (0.4) |
| | Diets: Low histamine diet | 83 (34) |
| | Diets: Low salt diet | 18 (7) |
| | Diets: Intermittent fasting | 95 (39) |
| | Diets: Other (please describe) | 55 (23) |
| | Plasmapheresis/Apheresis | 2 (1) |
| | Hyperbaric Oxygen Therapy (HBOT) | 15 (6) |

| | | |
|---|---|---|
| | Red light therapy | 27 (11) |
| | Lymphatic massage | 37 (15) |
| | Cranio-sacral massage | 24 (10) |
| | Home oxygen | 6 (2) |
| | Physical therapy: Vestibular | 15 (6) |
| | Physical therapy: Vestibular-ocular | 6 (2) |
| | Physical therapy: Speech | 4 (2) |
| | Physical therapy: Strengthening, stamina | 36 (15) |
| | Physical therapy: Rehabilitation | 35 (15) |
| | Vision therapy | 6 (2) |
| | Cognitive behavioral therapy (e.g. for tinnitus, hearing disorders) | 13 (5) |
| | Hearing devices/maskers for tinnitus | 11 (5) |
| | Integrative medicine treatments: IV ozone | 7 (3) |
| | Integrative medicine treatments: IV vitamins | 25 (10) |
| | Integrative medicine treatments: Treatment for EBV, CMV, herpes, Lymes disease | 9 (4) |
| | Chinese medicine treatments: Herbs | 25 (10) |
| | Chinese medicine treatments: Acupuncture | 60 (25) |
| | Ayurveda medicine | 7 (3) |
| | Lifestyle changes: Diet change | 95 (39) |
| | Lifestyle changes: No alcohol or caffeine | 105 (44) |
| | Lifestyle changes: Hydration, increase or decrease salt intake | 105 (44) |
| | Lifestyle changes: Limiting exercise or exertion | 124 (51) |
| | Chiropractic treatment | 44 (18) |

NSAID, Non-steroidal Anti-inflammatory Drug; PVS, Post-vaccination Syndrome
