## Supplemental Protocol for "Post-Vaccination Syndrome: A Descriptive Analysis of Reported Symptoms and Patient Experiences After Covid-19 Immunization"

### Appendix- Table of Contents

**A1 Protocol. Health status and symptom severity questions.**

1. Please choose one point in this 0-100 scale, which can best represent your health today (0 means the worst and 100 means the best). [“slider”]
2. We are trying to get a sense of how bad your vaccine injury symptoms are when you feel them the most. On the slider below, with 0 being a trivial illness and 100 being unbearable, please let us know what the worst days are like. ["slider"]

**A2 Protocol. Pre-pandemic comorbidities questions.**

Have you ever been told by a doctor before January 2020 that you have any of the following? Check all that apply

["multiple choice"]

1. Any allergies
2. Arthritis (including rheumatoid arthritis, gout, lupus, or fibromyalgia)
3. Asthma
4. Autoimmune disease (including lupus, scleroderma, etc.)
5. Bleeding disorder (including sickle cell disease or Thalassemia)
6. Blood clots
7. Cancer or malignancy of any kind
8. Cerebrovascular conditions affecting blood vessels to or in the brain (including stroke)
9. Chronic lung disease (including emphysema, chronic bronchitis, chronic obstructive pulmonary disease (COPD), or pulmonary fibrosis)
10. Cystic fibrosis
11. Diabetes
12. Ehlers Danlos Syndrome (hypermobile joints)
13. Gastrointestinal issues (including IBS or acid reflux)
14. Heart attack, also called myocardial infarction
15. Heart conditions (including coronary artery disease or cardiomyopathies)
16. Heart failure
17. High cholesterol
18. History of organ transplant (including kidney, liver, heart, or lung)
19. Hypertension or high blood pressure
20. Immunocompromised state (including weakened immune system from blood or bone marrow transplant, immune deficiencies, HIV, use of corticosteroids, or use of other immune-weakening medicines)
21. Kidney disease
22. Liver disease
23. Lyme disease
24. MCAS (mast cell activation syndrome) or other mast cell disorders
25. ME/CFS (myalgic encephalomyelitis/chronic fatigue syndrome)
26. Migraines
27. Neurologic conditions (including seizures, dementia, multiple sclerosis, Parkinson's, neuropathy, small fiber neuropathy, etc.)
28. Postural orthostatic hypotension (POTS) or dysautonomia
29. Spinal disorder(s)
30. Tremors/Internal vibrations
31. Other
32. None of the above

Have you ever been told by a doctor before January 2020 that you have any of the following? Check all that apply
["multiple choice"]

1. Depressive disorders
2. Anxiety disorders
3. Schizophrenia spectrum and other psychotic disorders
4. Bipolar and related disorders
5. Obsessive-compulsive and related disorders
6. Trauma- and stressor-related disorders
7. Feeding and eating disorders
8. Somatic symptoms (excessive thoughts, feelings and behaviors relating to the physical symptoms) and related disorders
9. Other
10. None of the above

**A3 Protocol. Current conditions questions.**

Currently, have you ever been told by a doctor that you have any of the following?
Check all that apply ["multiple choice"]

1. Any allergies
2. Arthritis (including rheumatoid arthritis, gout, lupus, or fibromyalgia)
3. Asthma
4. Autoimmune disease (including lupus, scleroderma, etc.)
5. Bleeding disorder (including sickle cell disease or Thalassemia)
6. Blood clots
7. Cancer or malignancy of any kind
8. Cerebrovascular conditions affecting blood vessels to or in the brain (including stroke)
9. Chronic lung disease (including emphysema, chronic bronchitis, chronic obstructive pulmonary disease (COPD), or pulmonary fibrosis)
10. Cystic fibrosis
11. Diabetes
12. Ehlers Danlos Syndrome (hypermobile joints)
13. Gastrointestinal issues (including IBS or acid reflux)
14. Heart attack, also called myocardial infarction
15. Heart conditions (including coronary artery disease or cardiomyopathies)
16. Heart failure
17. High cholesterol
18. History of organ transplant (including kidney, liver, heart, or lung)
19. Hypertension or high blood pressure
20. Immunocompromised state (including weakened immune system from blood or bone marrow transplant, immune deficiencies, HIV, use of corticosteroids, or use of other immune-weakening medicines)
21. Kidney disease
22. Liver disease
23. Long COVID
24. Lyme disease
25. MCAS (mast cell activation syndrome) or other mast cell disorders
26. ME/CFS (myalgic encephalomyelitis/chronic fatigue syndrome)
27. Migraines
28. Neurological conditions (including seizures, dementia, multiple sclerosis, Parkinson's, neuropathy, small fiber neuropathy, etc.)
29. Postural orthostatic hypotension (POTS) or dysautonomia
30. Spinal disorder(s)
31. Vaccine injury
32. Other
33. None of the above

Currently, have you ever been told by a doctor that you have any of the following?
Check all that apply ["multiple choice"]

1. Depressive disorders
2. Anxiety disorders
3. Schizophrenia spectrum and other psychotic disorders
4. Bipolar and related disorders
5. Obsessive-compulsive and related disorders
6. Trauma- and stressor-related disorders
7. Feeding and eating disorders
8. Somatic symptom and related disorders
9. Other
10. None of the above

**A4 Protocol. PVS symptoms questions.**

Please select all following health conditions that you have had as a result of vaccine injury. Check all that apply ["multiple choice"]

1. Abnormally low temperature
2. Fevers, including low-grade fevers
3. Chills but no fever
4. Heat intolerance
5. Cold intolerance
6. Night sweats
7. Other
8. None of the above

Please select all following health conditions that you have had as a result of vaccine injury. Check all that apply ["multiple choice"]

1. Trouble falling or staying asleep
2. Sleeping more than usual
3. Nightmares
4. Exercise intolerance
5. Excessive fatigue
6. Other
7. None of the above

Please select all following health conditions that you have had as a result of vaccine injury. Check all that apply ["multiple choice"]

1. Burning sensations
2. Tremors or shakiness
3. Internal tremors or buzzing/vibration
4. Tingling, pins and needles, numbness
5. Neuropathy (nerve sensations including pain) anywhere in the body
6. Seizures
7. Other
8. None of the above

Please select all following health conditions that you have had as a result of vaccine injury. Check all that apply ["multiple choice"]

1. Abdominal pain
2. Acid reflux or heartburn
3. Diarrhea
4. Constipation
5. Nausea/Vomiting
6. Loss of appetite
7. Other
8. None of the above

Please select all following health conditions that you have had as a result of vaccine injury. Check all that apply ["multiple choice"]

1. Sore throat
2. Congested or runny nose
3. Palpitations (improper beating of the heart due to electrical impulse problems)
4. Bilateral neck throbbing around lymph nodes

5. Costochondritis (pain in the cartilage that connects a rib to the breastbone)
6. Cough
7. Coughing up blood
8. Cold or burning feeling in lungs
9. Difficulty swallowing
10. Throat pain or discomfort
11. Lump in throat
12. Phlegm in back of throat
13. Postnasal drip
14. Runny nose
15. Swollen lymph nodes
16. Tachycardia (rapid heartbeat) at rest
17. Tachycardia (rapid heartbeat) after standing up
18. Wheezing
19. Shortness of breath or difficulty breathing
20. Other
21. None of the above

Please select all following health conditions that you have had as a result of vaccine injury.
Check all that apply ["multiple choice"]

1. Bone aches
2. Migraine
3. Headache
4. Calf cramps
5. Pressure at base of head
6. Jaw pain
7. Joint pain
8. Kidney pain
9. Mouth sores or sore tongue
10. Muscle or body aches
11. Persistent chest pain or pressure
12. Painful scalp
13. Sharp or sudden chest pain
14. Other
15. None of the above

Please select all following health conditions that you have had as a result of vaccine injury.
Check all that apply ["multiple choice"]

1. Changed sense of taste
2. Changed sense of smell
3. Floaters or flashes of light in vision
4. Loss of hearing
5. Loss or decrease in quality of vision/blurry vision
6. Phantom smells
7. Phantom tastes
8. Hallucinations (visual or auditory)
9. Tinnitus or humming in ears
10. Other

11. None of the above

Please select all following health conditions that you have had as a result of vaccine injury.
Check all that apply ["multiple choice"]

1. Skin bruising
2. Change in nails (i.e. white spots, brittleness, change in moons)
3. Tender or itchy rash or chilblains on the toes or foot)
4. Cracked or dry lips
5. Dental problems (e.g., chipped tooth, tooth loss)
6. Discoloration of the skin (for example: purple or blue on the hands or feet, no blistering)
7. Dry or peeling skin
8. Dry scalp or dandruff
9. Hair loss
10. Itchiness
11. Tender or itchy rash not on foot
12. Chilblains (itching, bumps, red- to violet-colored patches on the hands or feet)
13. Other
14. None of the above

Please select all following health conditions that you have had as a result of vaccine injury.
Check all that apply ["multiple choice"]

1. Constant thirst
2. Changes in voice
3. Clogged ears
4. Dizziness
5. Dry eyes
6. Fatigue
7. Irregular or skipped menstrual cycles
8. Menstrual cycles that are heavier or lighter than normal
9. New allergies
10. Inability to eat or tolerate food
11. Swollen hands or feet
12. Fainting
13. Weakened neck
14. Other
15. None of the above

Please select all that you have. Check all that apply ["multiple choice"]

1. Anxiety
2. Confusion
3. Brain fog; difficulty concentrating or focusing
4. Feelings of impending doom
5. Memory problems
6. Difficulty speaking properly
7. Suicidal thoughts
8. Other
9. None of the above
